# Molecular Architecture of Early Dissemination and Massive Second Wave of the SARS-CoV-2 Virus in a Major Metropolitan Area

**DOI:** 10.1101/2020.09.22.20199125

**Authors:** S. Wesley Long, Randall J. Olsen, Paul A. Christensen, David W. Bernard, James J. Davis, Maulik Shukla, Marcus Nguyen, Matthew Ojeda Saavedra, Prasanti Yerramilli, Layne Pruitt, Sishir Subedi, Hung-Che Kuo, Heather Hendrickson, Ghazaleh Eskandari, Hoang A.T. Nguyen, J. Hunter Long, Muthiah Kumaraswami, Jule Goike, Daniel Boutz, Jimmy Gollihar, Jason S. McLellan, Chia-Wei Chou, Kamyab Javanmardi, Ilya J. Finkelstein, James M. Musser

**Author notes:** Address correspondence to: James M. Musser, M.D., Ph.D., Department of Pathology and Genomic Medicine, Houston Methodist Research Institute, 6565 Fannin Street, Suite B490, Houston, Texas 77030. S.W.L., R.J.O., and P.A.C. contributed equally to this article. The order of co-first authors was determined by discussion and mutual agreement between the three co-first authors. This article is a direct contribution from James M. Musser, a Fellow of the American Academy of Microbiology, who arranged for and secured reviews by Barry N. Kreiswirth, Center for Discovery and Innovation, Hackensack Meridian Health, New Jersey; and David M. Morens, National Institute of Allergy and Infectious Diseases, National Institutes of Health, Maryland.

## Abstract

We sequenced the genomes of 5,085 SARS-CoV-2 strains causing two COVID-19 disease waves in metropolitan Houston, Texas, an ethnically diverse region with seven million residents. The genomes were from viruses recovered in the earliest recognized phase of the pandemic in Houston, and an ongoing massive second wave of infections. The virus was originally introduced into Houston many times independently. Virtually all strains in the second wave have a Gly614 amino acid replacement in the spike protein, a polymorphism that has been linked to increased transmission and infectivity. Patients infected with the Gly614 variant strains had significantly higher virus loads in the nasopharynx on initial diagnosis. We found little evidence of a significant relationship between virus genotypes and altered virulence, stressing the linkage between disease severity, underlying medical conditions, and host genetics. Some regions of the spike protein - the primary target of global vaccine efforts - are replete with amino acid replacements, perhaps indicating the action of selection. We exploited the genomic data to generate defined single amino acid replacements in the receptor binding domain of spike protein that, importantly, produced decreased recognition by the neutralizing monoclonal antibody CR30022. Our study is the first analysis of the molecular architecture of SARS-CoV-2 in two infection waves in a major metropolitan region. The findings will help us to understand the origin, composition, and trajectory of future infection waves, and the potential effect of the host immune response and therapeutic maneuvers on SARS-CoV-2 evolution.

**IMPORTANCE:** There is concern about second and subsequent waves of COVID-19 caused by the SARS-CoV-2 coronavirus occurring in communities globally that had an initial disease wave. Metropolitan Houston, Texas, with a population of 7 million, is experiencing a massive second disease wave that began in late May 2020. To understand SARS-CoV-2 molecular population genomic architecture, evolution, and relationship between virus genotypes and patient features, we sequenced the genomes of 5,085 SARS-CoV-2 strains from these two waves. Our study provides the first molecular characterization of SARS-CoV-2 strains causing two distinct COVID-19 disease waves.

## [Introduction]

**P**andemic disease caused by the severe acute respiratory syndrome coronavirus 2 (SARS-CoV-2) virus is now responsible for massive human morbidity and mortality worldwide (1-5). The virus was first documented to cause severe respiratory infections in Wuhan, China, beginning in late December 2019 (6-9). Global dissemination occurred extremely rapidly and has affected major population centers on most continents (10, 11). In the United States, the Seattle and the New York City (NYC) regions have been especially important centers of COVID-19 disease caused by SARS-CoV-2. For example, as of August 19, 2020, there were 227,419 confirmed SARS-CoV-2 cases in NYC, causing 56,831 hospitalizations and 19,005 confirmed fatalities and 4,638 probable fatalities (12). Similarly, in Seattle and King County, 17,989 positive patients and 696 deaths have been reported as of August 18, 2020 (13).

The Houston metropolitan area is the fourth largest and most ethnically diverse city in the United States, with a population of approximately 7 million (14, 15). The 2,400-bed Houston Methodist health system has seven hospitals and serves a large, multiethnic, and socioeconomically diverse patient population throughout greater Houston (13, 14). The first COVID-19 case in metropolitan Houston was reported on March 5, 2020 with community spread occurring one week later (16). Many of the first cases in our region were associated with national or international travel in areas known to have SARS-CoV-2 virus outbreaks (16). A central molecular diagnostic laboratory serving all Houston Methodist hospitals and our very early adoption of a molecular test for the SARS-CoV-2 virus permitted us to rapidly identify positive patients and interrogate genomic variation among strains causing early infections in the greater Houston area. Our analysis of SARS-CoV-2 genomes causing disease in Houston has continued unabated since early March and is ongoing. Genome sequencing and related efforts were expanded extensively in late May as we recognized that a prominent second wave was underway (**Figure 1**).

**FIG 1.**
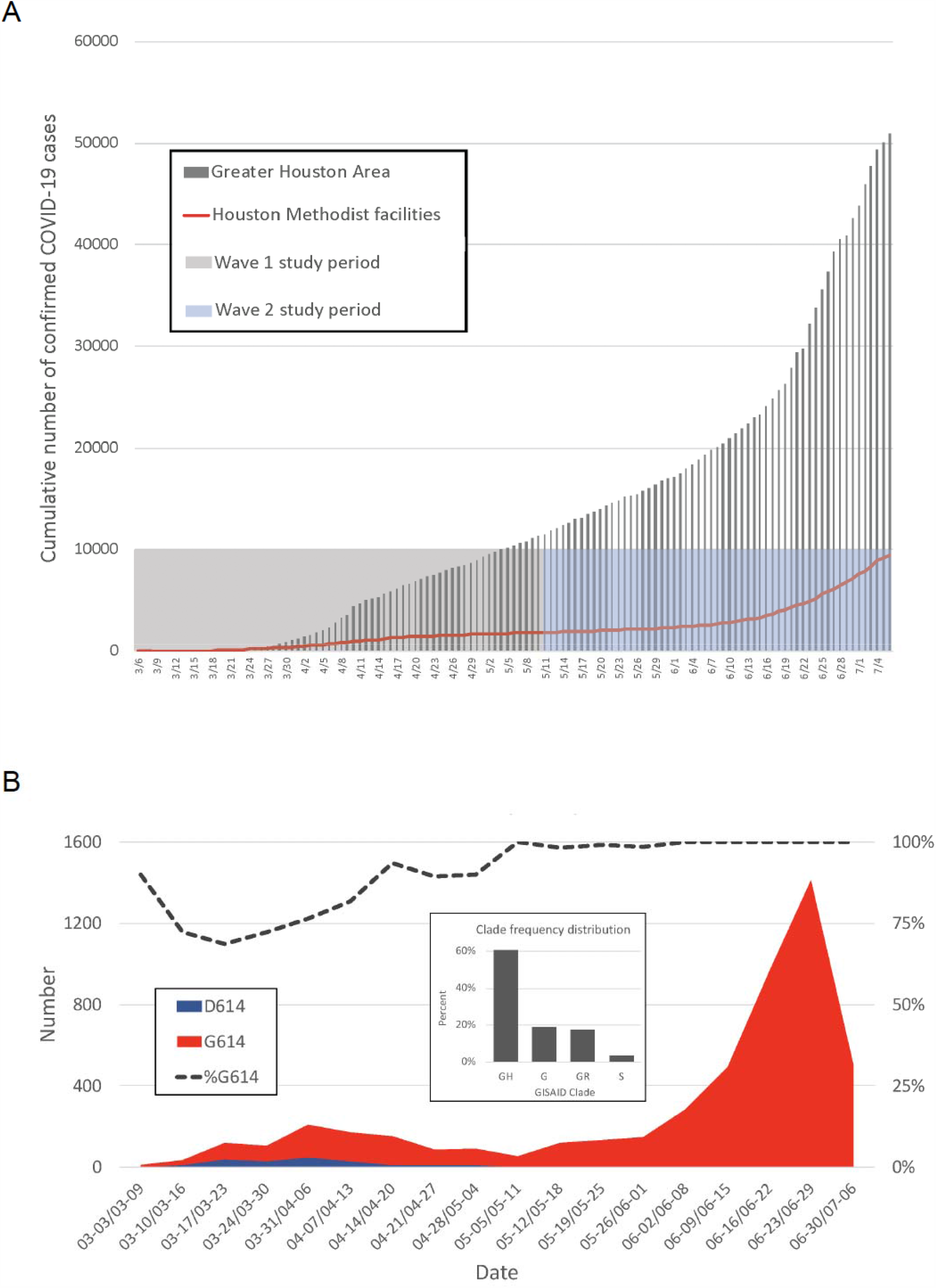
(A) Confirmed COVID-19 cases in the Greater Houston Metropolitan region. Cumulative number of COVID-19 patients over time through July 7, 2020. Counties include Austin, Brazoria, Chambers, Fort Bend, Galveston, Harris, Liberty, Montgomery, and Waller. The shaded area represents the time period during which virus genomes characterized in this study were recovered from COVID-19 patients. The red line represents the number of COVID-19 patients diagnosed in the Houston Methodist Hospital Molecular Diagnostic Laboratory. (B) Distribution of strains with either the Asp614 or Gly614 amino acid variant in spike protein among the two waves of COVID-19 patients diagnosed in the Houston Methodist Hospital Molecular Diagnostic Laboratory. The large inset shows major clade frequency for the time frame studied.

Here, we report that SARS-CoV-2 was introduced to the Houston area many times, independently, from diverse geographic regions, with virus genotypes representing genetic clades causing disease in Europe, Asia, South America and elsewhere in the United States. There was widespread community dissemination soon after COVID-19 cases were reported in Houston. Strains with a Gly614 amino acid replacement in the spike protein, a polymorphism that has been linked to increased transmission and *in vitro* cell infectivity, increased significantly over time and caused virtually all COVID-19 cases in the massive second disease wave. Patients infected with strains with the Gly614 variant had significantly higher virus loads in the nasopharynx on initial diagnosis. Some naturally occurring single amino acid replacements in the receptor binding domain (RBD) of spike protein resulted in decreased reactivity with a neutralizing monoclonal antibody, consistent with the idea that some virus variants arise due to host immune pressure.

## RESULTS

### Description of metropolitan Houston

Houston, Texas, is located in the southwestern United States, 50 miles inland from the Gulf of Mexico. It is the most ethnically diverse city in the United States (14). Metropolitan Houston is comprised predominantly of Harris County plus parts of eight contiguous surrounding counties. In the aggregate, the metropolitan area includes 9,444 square miles. The estimated population size of metropolitan Houston is 7 million (https://www.houston.org/houston-data).

### Epidemic curve characteristics over two disease waves

The first confirmed case of COVID-19 in the Houston metropolitan region was reported on March 5, 2020 (16), and the first confirmed case diagnosed in Houston Methodist hospitals was reported on March 6, 2020. The epidemic curve indicated a first wave of COVID-19 cases that peaked around April 11-15, followed by a decline in cases until May 11. Soon thereafter, the slope of the case curve increased with a very sharp uptick in confirmed cases beginning on June 12 (**Figure 1B**). We consider May 11 as the transition between waves, as this date is the inflection point of the cumulative new cases curve and had the absolute lowest number of new cases in the mid-May time period. Thus, for the data presented herein, wave 1 is defined as March 5 through May 11, 2020, and wave 2 is defined as May 12 through July 7, 2020. Epidemiologic trends within the Houston Methodist Hospital population were mirrored by data from Harris County and the greater metropolitan Houston region (**Figure 1A**). Through the 7^th^ of July, 25,366 COVID-19 cases were reported in Houston, 37,776 cases in Harris County, and 53,330 in metropolitan Houston, including 9,823 cases in Houston Methodist facilities (inpatients and outpatients) (https://www.tmc.edu/coronavirusupdates/infection-rate-in-the-greater-houston-area/ and https://harriscounty.maps.arcgis.com/apps/opsdashboard/index.html#/c0de71f8ea484b85bb5efcb7c07c6914).

During the first wave (early March through May 11), 11,476 COVID-19 cases were reported in Houston, including 1,729 cases in the Houston Methodist Hospital system. Early in the first wave (from March 5 through March 30, 2020), we tested 3,080 patient specimens. Of these, 406 (13.2%) samples were positive for SARS-CoV-2, representing 40% (358/898) of all confirmed cases in metropolitan Houston during that time period. As our laboratory was the first hospital-based facility to have molecular testing capacity for SARS-CoV-2 available on site, our strain samples are likely representative of COVID-19 infections during the first wave.

For the entire study period (March 5 through July 7, 2020), we tested 68,418 specimens from 55,800 patients. Of these, 9,121 patients (16.4%) had a positive test result, representing 17.1% (9,121/53,300) of all confirmed cases in metropolitan Houston. Thus, our strain samples are also representative of those responsible for COVID-19 infections in the massive second wave.

To test the hypothesis that, on average, the two waves affected different groups of patients, we analyzed individual patient characteristics (hospitalized and non-hospitalized) in each wave. Consistent with this hypothesis, we found significant differences in the COVID-19 patients in each wave (**Table S1**). For example, patients in the second wave were significantly younger, had fewer comorbidities, were more likely to be Hispanic/Latino (by self-report), and lived in zip codes with lower median incomes (**Table S1**). A detailed analysis of the characteristics of patients hospitalized in Houston Methodist facilities in the two waves has recently been published (17).

### SARS-CoV-2 genome sequencing and phylogenetic analysis

To investigate the genomic architecture of the virus across the two waves, we sequenced the genomes of 5,085 SARS-CoV-2 strains dating to the earliest time of confirmed COVID-19 cases in Houston. Analysis of SARS-CoV-2 strains causing disease in the first wave (March 5 through May 11) identified the presence of many diverse virus genomes that, in the aggregate, represent the major clades identified globally to date (**Figure 1B**). Clades G, GH, GR, and S were the four most abundantly represented phylogenetic groups (**Figure 1B**).Strains with the Gly614 amino acid variant in spike protein represented 82% of the SARS-CoV-2 strains in wave 1, and 99.9% in wave 2 (*p*<0.0001; Fisher’s exact test) (**Figure 1B**). This spike protein variant is characteristic of clades G, GH, and GR. Importantly, strains with the Gly614 variant represented only 71% of the specimens sequenced in March, the early part of wave 1 (**Figure 1B**). We attribute the decrease in strains with this variant observed in the first two weeks of March (**Figure 1B**) to fluctuation caused by the relatively fewer COVID-19 cases occurring during this period.

### Relating spatiotemporal genome analysis with virus genotypes over two disease waves

We examined the spatial and temporal mapping of genomic data to investigate community spread during wave 1 (**Figure 2**). Rapid and widespread community dissemination occurred soon after the initial COVID-19 cases were reported in Houston. The heterogenous virus genotypes present very early in wave 1 indicate that multiple strains independently entered metropolitan Houston, rather than introduction and spread of a single strain. An important observation was that strains of most of the individual subclades were distributed over broad geographic areas (**Figure S1**). These findings are consistent with the known ability of SARS-CoV-2 to spread very rapidly from person to person.

**FIG 2.**
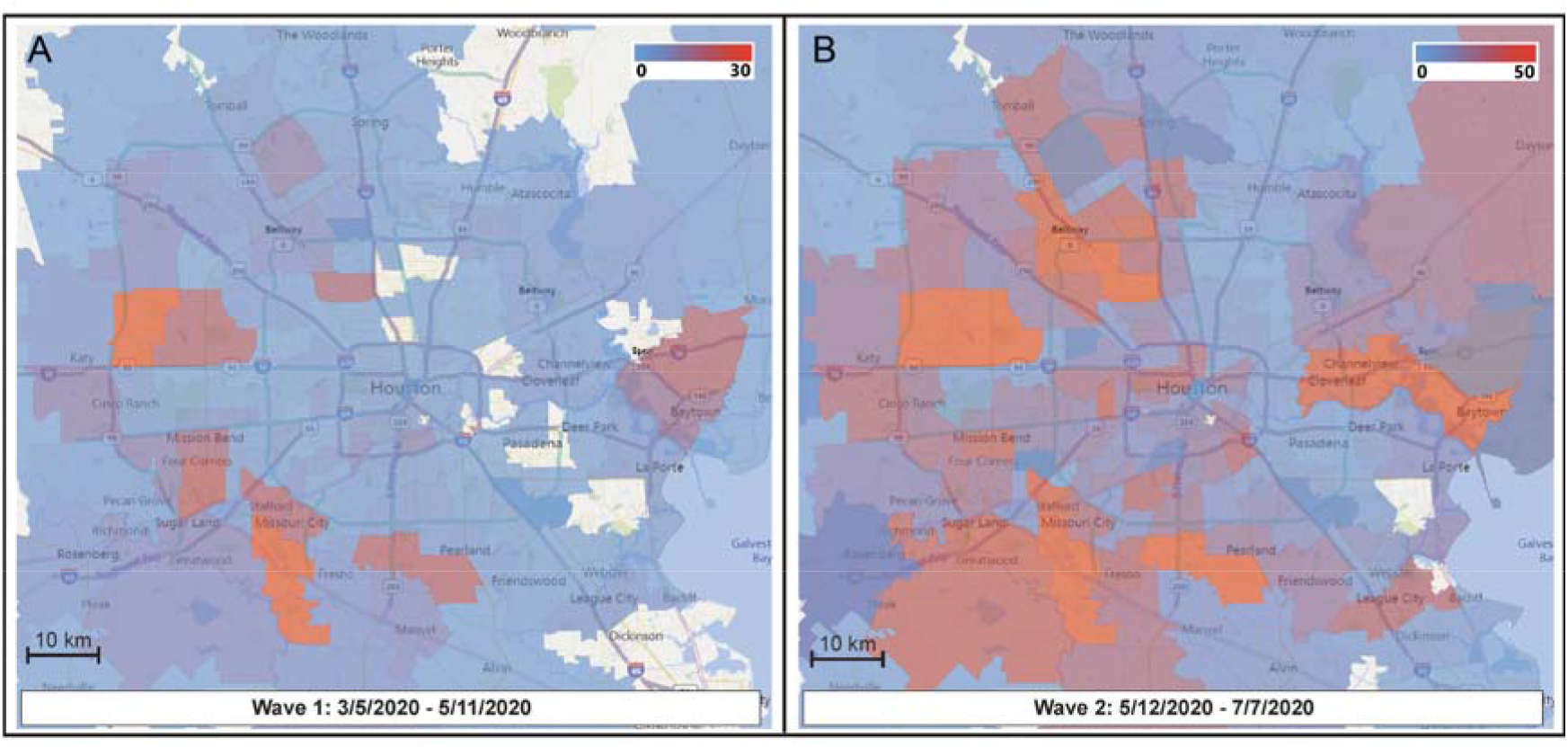
Sequential time-series heatmaps for all COVID-19 Houston Methodist patients during the study period. Geospatial distribution of COVID-19 patients is based on zip code. Panel A (left) shows geospatial distribution of sequenced SARS-CoV-2 strains in wave 1 and panel B (right) shows wave 2 distribution. The collection dates are shown at the bottom of each panel. The insets refer to numbers of strains in the color spectrum used. Note difference in numbers of strains used in panel A and panel B insets.

### Relationship between virus clades, clinical characteristics of infected patients, and additional metadata

It is possible that SARS-CoV-2 genome subtypes have different clinical characteristics, analogous to what is believed to have occurred with Ebola virus (18-20) and known to occur for other pathogenic microbes (21). As an initial examination of this issue in SARS-CoV-2, we tested the hypothesis that patients with disease severe enough to warrant hospitalization were infected with a non-random subset of virus genotypes. We also examined the association between virus clades and disease severity based on overall mortality, highest level of required care (intensive care unit, intermediate care unit, inpatient or outpatient), need for mechanical ventilation, and length of stay. There was no simple relationship between virus clades and disease severity using these four indicators. Similarly, there was no simple relationship between virus clades and other metadata, such as sex, age, or ethnicity (**Figure S2**).

### Machine learning analysis

Machine learning models can be used to identify complex relationships not revealed by statistical analyses. We built machine learning models to test the hypothesis that virus genome sequence can predict patient outcomes including mortality, length of stay, level of care, ICU admission, supplemental oxygen use, and mechanical ventilation. Models to predict outcomes based on virus genome sequence alone resulted in low F1 scores less than 50% (0.41 – 0.49) and regression models showed similarly low R^2^ values (−0.01 – -0.20) (**Table S2**). F1 scores near 50% are indicative of classifiers that are performing similarly to random chance. The use of patient metadata alone to predict patient outcome improved the model’s F1 scores by 5-10% (0.51 – 0.56) overall. The inclusion of patient metadata with virus genome sequence data improved most predictions of outcomes, compared to genome sequence alone, to 50% to 55% F1 overall (0.42 – 0.55) in the models (**Table S2**). The findings are indicative of two possibilities that are not mutually exclusive. First, patient metadata, such as age and sex, may provide more signal for the model to use and thus result in better accuracies. Second, the model’s use of single nucleotide polymorphisms (SNPs) may have resulted in overfitting. Most importantly, no SNP predicted a significant difference in outcome. A table of classifier accuracy scores and performance information is provided in **Table S2**.

### Patient outcome and metadata correlations

Overall, very few metadata categories correlated with patient outcomes (**Table S3**). Mortality was independently correlated with increasing age, with a Pearson correlation coefficient (PCC) equal to 0.27. This means that 27% of the variation in mortality can be predicted from patient age. Length of stay correlated independently with increasing age (PCC=0.20). All other patient metadata correlations to outcomes had PCC less than 0.20 (**Table S3**).

We further analyzed outcomes correlated to isolates from wave 1 and 2, and the presence of the Gly614 variant in spike protein. Being in wave 1 was independently correlated with mechanical ventilation days, overall length of stay, and ICU length of stay, with PCC equal to 0.20, 0.18, and 0.14, respectively. Importantly, the presence of the Gly614 variant did not correlate with patient outcomes (**Table S3**).

### Analysis of the *nsp12* polymerase gene

The SARS-CoV-2 genome encodes an RNA-dependent RNA polymerase (RdRp, also referred to as Nsp12) used in virus replication (22-25). Two amino acid substitutions (Phe479Leu and Val556Leu) in RdRp each confer significant resistance *in vitro* to remdesivir, an adenosine analog (26). Remdesivir is inserted into RNA chains by RdRp during replication, resulting in premature termination of RNA synthesis and inhibition of virus replication. This compound has shown prophylactic and therapeutic benefit against MERS-CoV and SARS-CoV-2 experimental infection in rhesus macaques (27, 28). Recent reports indicate that remdesivir has therapeutic benefit in some COVID-19 hospitalized patients (29-33), leading it to be now widely used in patients worldwide. Thus, it may be important to understand variation in RdRp in large strain samples.

To acquire data about allelic variation in the *nsp12* gene, we analyzed our 5,085 virus genomes. The analysis identified 265 SNPs, including 140 nonsynonymous (amino acid-altering) SNPs, resulting in amino acid replacements throughout the protein (**Table 1, Figure 3, Figure 4, Figure S3**, and **Figure S4**). The most common amino acid change was Pro322Leu, identified in 4,893 of the 5,085 (96%) patient isolates. This amino acid replacement is common in genomes from clades G, GH, and GR, which are distinguished from other SARS-CoV-2 clades by the presence of the Gly614 amino acid change in the spike protein. Most of the other amino acid changes in RdRp were present in relatively small numbers of strains, and some have been identified in other isolates in a publicly available database (34). Five prominent exceptions included amino acid replacements: Ala15Val in 138 strains, Met462Ile in 59 strains, Met600Ile in 75 strains, Thr907Ile in 45 strains, and Pro917Ser in 80 strains. All 75 Met600Ile strains were phylogenetically closely related members of clade G, and also had the Pro322Leu amino acid replacement characteristic of this clade (**Figure S3**). These data indicate that the Met600Ile change is likely the evolved state, derived from a precursor strain with the Pro322Leu replacement. Similarly, we investigated phylogenetic relationships among strains with the other four amino acid changes noted above. In all cases, the vast majority of strains with each amino acid replacement were found among individual subclades of strains (**Figure S3**).

**Table 1.** Nonsynonymous SNPs of SARS-CoV-2 nsp12. 1496

| **Genomic Locus** | **Gene Locus** | **Amino Acid Change** | **Domain** | **Wave 1 (n=1026)** | **Wave 2 (n=4059)** | **Total (n=5085)** |
| --- | --- | --- | --- | --- | --- | --- |
| 13446 | C3T | A1V | N-terminus |  | 2 | 2 |
| 13448 | G5A | D2N | N-terminus | 1 |  | 1 |
| 13487 | C44T | A15V | N-terminus |  | 138 | 138 |
| 13501 | C58T | P20S | N-terminus |  | 1 | 1 |
| 13514 | G71A | G24D | N-terminus |  | 3 | 3 |
| 13517 | C74T | T25I | N-terminus |  | 4 | 4 |
| 13520 | G77A | S26N | N-terminus |  | 1 | 1 |
| 13523 | C80T | T27I | N-terminus |  | 1 | 1 |
| 13526 | A83C | D28A | N-terminus |  | 1 | 1 |
| 13564 | G121A | V41I | B hairpin |  | 1 | 1 |
| 13568 | C125T | A42V | B hairpin | 1 |  | 1 |
| 13571 | G128T | G43V | B hairpin | 1 |  | 1 |
| 13576 | G133T | A45S | B hairpin |  | 12 | 12 |
| 13617 | G174T | K58N | NiRAN |  | 1 | 1 |
| 13618 | G175T | D59Y | NiRAN |  | 24 | 24 |
| 13620 | C177G | D59E | NiRAN |  | 1 | 1 |
| 13627 | G184T | D62Y | NiRAN | 1 |  | 1 |
| 13661 | G218A | R73K | NiRAN |  | 1 | 1 |
| 13667 | C224T | T75I | NiRAN |  | 2 | 2 |
| 13694 | C251T | T84I | NiRAN |  | 1 | 1 |
| 13712 | A269G | K90R | NiRAN |  | 1 | 1 |
| 13726 | G283A | V95I | NiRAN |  | 1 | 1 |
| 13730 | C287T | A96V | NiRAN | 2 | 2 | 4 |
| 13762 | G319C | G107R | NiRAN |  | 1 | 1 |
| 13774 | C331A | P111T | NiRAN |  | 1 | 1 |
| 13774 | C331T | P111S | NiRAN |  | 15 | 15 |
| 13777 | C334T | H112Y | NiRAN |  | 1 | 1 |
| 13790 | A347G | Q116R | NiRAN |  | 2 | 2 |
| 13835 | G392T | R131M | NiRAN |  | 1 | 1 |
| 13858 | G415T | D139Y | NiRAN |  | 3 | 3 |
| 13862 | C419T | T140I | NiRAN | 1 | 5 | 6 |
| 13868 | A425G | K142R | NiRAN | 1 |  | 1 |
| 13897 | G454T | D152Y | NiRAN |  | 4 | 4 |
| 13901 | A458G | D153G | NiRAN |  | 2 | 2 |
| 13957 | C514T | R172C | NiRAN |  | 2 | 2 |
| 13963 | T520C | Y174H | NiRAN |  | 1 | 1 |
| 13966 | G523A | A175T | NiRAN |  | 1 | 1 |
| 13975 | G532T | G178C | NiRAN |  | 4 | 4 |
| 13984 | G541A | V181I | NiRAN |  | 1 | 1 |
| 13994 | C551T | A184V | NiRAN |  | 8 | 8 |
| 14104 | T661C | F221L | NiRAN | 2 |  | 2 |
| 14109 | A666G | I222M | NiRAN | 1 |  | 1 |
| 14120 | C677T | P226L | NiRAN |  | 2 | 2 |
| 14185 | A742G | R248G | NiRAN |  | 1 | 1 |
| 14187 | G744T | R248S | NiRAN |  | 1 | 1 |
| 14188 | G745A | A249T | NiRAN |  | 1 | 1 |
| 14225 | C782A | T261K | Interface |  | 4 | 4 |
| 14230 | C787T | P263S | Interface |  | 1 | 1 |
| 14233 | T790C | Y264H | Interface |  | 1 | 1 |
| 14241 | G798T | K266N | Interface |  | 1 | 1 |
| 14290 | G847T | D283Y | Interface |  | 1 | 1 |
| 14335 | G892T | V298F | Interface |  | 8 | 8 |
| 14362 | C919A | L307M | Interface |  | 2 | 2 |
| 14371 | G928C | A310P | Interface | 1 |  | 1 |
| 14396 | C953T | T318I | Interface |  | 1 | 1 |
| 14398 | G955T | V319L | Interface |  | 1 | 1 |
| 14407 | C964T | P322S | Interface |  | 2 | 2 |
| 14408 | C965T | P322L | Interface | 843 | 4050 | 4893 |
| 14500 | G1057T | V353L | Interface |  | 5 | 5 |
| 14536 | C1093T | L365F | Interface |  | 1 | 1 |
| 14557 | G1114T | V372L | Fingers |  | 4 | 4 |
| 14584 | G1141T | A381S |  |  | 1 | 1 |
| 14585 | C1142T | A381V | Fingers |  | 10 | 10 |
| 14593 | G1150A | G384S | Fingers | 1 |  | 1 |
| 14657 | C1214T | A405V | Fingers |  | 1 | 1 |
| 14708 | C1265T | A422V |  |  | 1 | 1 |
| 14747 | A1304G | E435G | Fingers |  | 2 | 2 |
| 14768 | C1325T | A442V | Fingers |  | 21 | 21 |
| 14786 | C1343T | A448V | Fingers | 3 | 6 | 9 |
| 14821 | C1378T | P460S |  |  | 1 | 1 |
| 14829 | G1386T | M462I | Fingers |  | 59 | 59 |
| 14831 | G1388T | C463F | Fingers |  | 3 | 3 |
| 14857 | G1414T | V472F |  |  | 1 | 1 |
| 14870 | A1427G | D476G | Fingers |  | 5 | 5 |
| 14874 | G1431T | K477N | Fingers | 1 |  | 1 |
| 14912 | A1469G | N490S | Fingers | 1 | 1 | 2 |
| 14923 | G1480A | V494I | Fingers |  | 2 | 2 |
| 14980 | C1537T | L513F | Fingers | 1 | 1 | 2 |
| 14990 | A1547G | D516G |  |  | 1 | 1 |
| 15006 | G1563C | E521D | Fingers | 2 | 3 | 5 |
| 15016 | G1573T | A525S | Fingers |  | 3 | 3 |
| 15026 | C1583T | A528V | Fingers | 5 | 1 | 6 |
| 15037 | C1594T | R532C | Fingers |  | 1 | 1 |
| 15100 | G1657C | A553P | Fingers |  | 1 | 1 |
| 15101 | C1658T | A553V | Fingers |  | 1 | 1 |
| 15124 | A1681G | I561V | Fingers |  | 2 | 2 |
| 15202 | G1759C | V587L | Palm |  | 7 | 7 |
| 15211 | A1768G | T590A |  |  | 1 | 1 |
| 15226 | G1783A | G595S | Palm |  | 1 | 1 |
| 15243 | G1800T | M600I | Palm | 71 | 4 | 75 |
| 15251 | C1808G | T603S | Palm |  | 1 | 1 |
| 15257 | A1814G | Y605C |  |  | 1 | 1 |
| 15260 | G1817A | S606N | Palm |  | 1 | 1 |
| 15327 | G1884T | M628I | Fingers | 3 | 1 | 4 |
| 15328 | C1885T | L629F | Fingers | 1 |  | 1 |
| 15334 | A1891G | I631V | Fingers |  | 1 | 1 |
| 15341 | C1898T | A633V | Fingers |  | 1 | 1 |
| 15352 | C1909T | L637F | Fingers |  | 1 | 1 |
| 15358 | C1915T | R639C | Fingers |  | 1 | 1 |
| 15362 | A1919G | K640R | Fingers |  | 1 | 1 |
| 15364 | C1921G | H641D | Fingers | 1 |  | 1 |
| 15368 | C1925T | T642I | Fingers | 1 |  | 1 |
| 15380 | G1937T | S646I | Fingers | 1 |  | 1 |
| 15386 | C1943T | S648L | Fingers |  | 2 | 2 |
| 15391 | C1948T | R650C | Fingers |  | 1 | 1 |
| 15406 | G1963T | A655S | Fingers |  | 3 | 3 |
| 15407 | C1964T | A655V | Fingers | 1 |  | 1 |
| 15436 | A1993G | M665V | Fingers |  | 2 | 2 |
| 15438 | G1995T | M665I | Fingers |  | 24 | 24 |
| 15452 | G2009T | G670V | Fingers |  | 28 | 28 |
| 15487 | G2044C | G682R | Palm |  | 1 | 1 |
| 15497 | C2054A | T685K | Palm |  | 1 | 1 |
| 15572 | A2129G | D710G | Palm |  | 1 | 1 |
| 15596 | A2153G | Y718S | Palm | 2 |  | 2 |
| 15619 | C2176T | L726F | Palm | 1 |  | 1 |
| 15638 | G2195A | R732K | Palm | 1 |  | 1 |
| 15640 | A2197G | N733D | Palm | 1 |  | 1 |
| 15640 | A2197T | N733Y | Palm | 1 |  | 1 |
| 15655 | A2212G | T738A | Palm |  | 2 | 2 |
| 15656 | C2213T | T738I | Palm |  | 2 | 2 |
| 15658 | G2215A | D739N | Palm |  | 2 | 2 |
| 15664 | G2221A | V741M | Palm |  | 1 | 1 |
| 15715 | T2272C | S758P | Palm |  | 1 | 1 |
| 15760 | G2317A | G773S | Palm | 1 |  | 1 |
| 15761 | G2318A | G773D | Palm |  | 1 | 1 |
| 15827 | A2384G | E795G | Palm | 1 |  | 1 |
| 15848 | C2405T | T802I | Palm |  | 1 | 1 |
| 15850 | G2407T | D803Y | Palm |  | 1 | 1 |
| 15853 | C2410T | L804F | Palm |  | 2 | 2 |
| 15878 | G2435T | C812F | Palm |  | 1 | 1 |
| 15886 | C2443T | H815Y | Palm |  | 1 | 1 |
| 15906 | G2463T | Q821H | Thumb | 1 | 1 | 2 |
| 15908 | G2465T | G822V | Thumb |  | 1 | 1 |
| 15979 | A2536G | I846V | Thumb | 4 |  | 4 |
| 16045 | C2602T | L868F | Thumb |  | 1 | 1 |
| 16084 | C2641T | H881Y | Thumb |  | 1 | 1 |
| 16148 | A2705G | Y902C | Thumb |  | 1 | 1 |
| 16163 | C2720T | T907I | Thumb |  | 45 | 45 |
| 16178 | C2735T | S912L | Thumb |  | 2 | 2 |
| 16192 | C2749T | P917S | Thumb |  | 80 | 80 |

**FIG 3.**
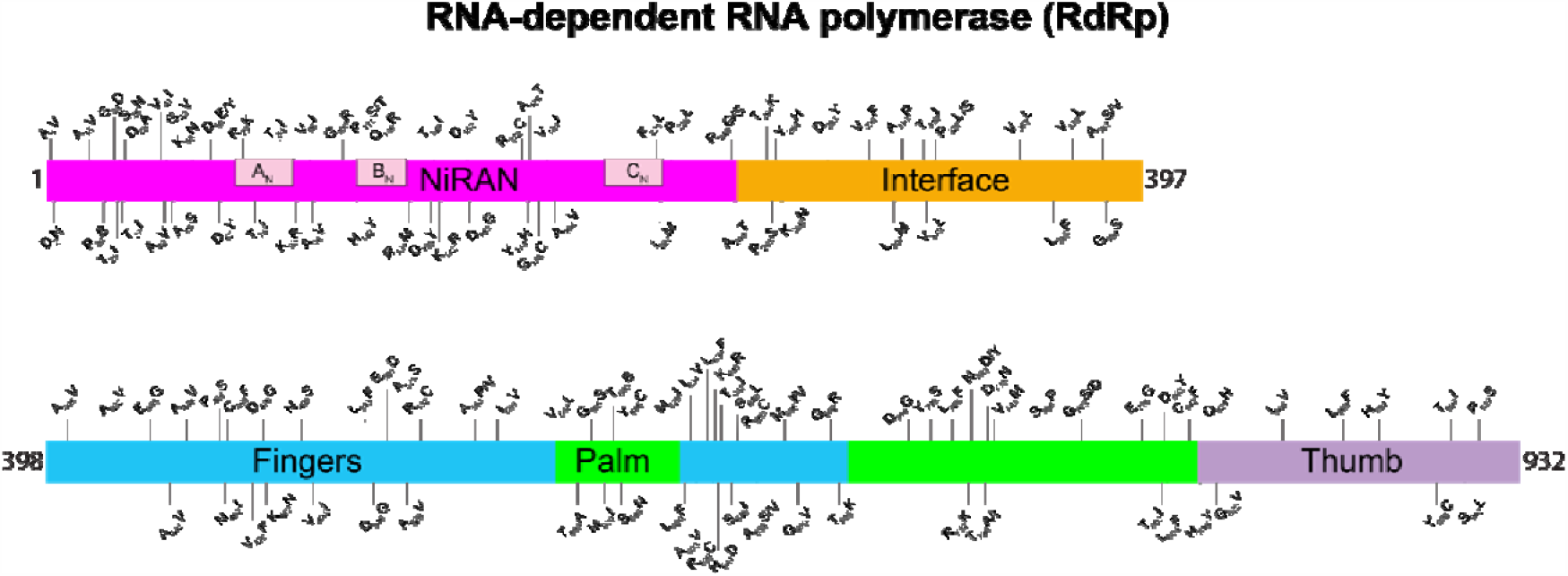
Location of amino acid replacements in RNA-dependent RNA polymerase (RdRp/Nsp12) among the 5,085 genomes of SARS-CoV-2 sequenced. The various RdRp domains are color-coded. The numbers refer to amino acid site. Note that several amino acid sites have multiple variants identified.

**FIG 4.**
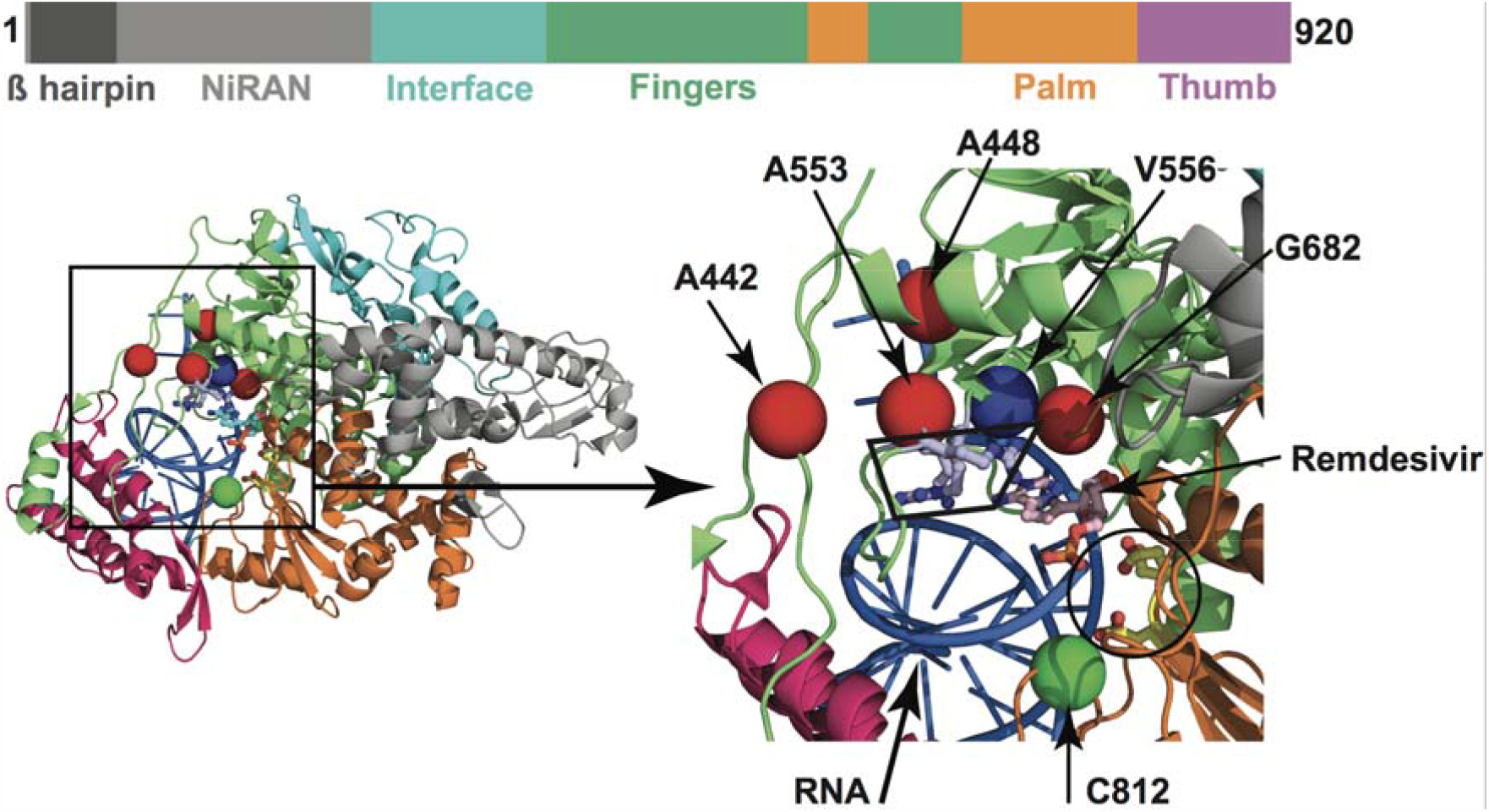
Amino acid changes identified in Nsp12 (RdRp) in this study that may influence interaction with remdesivir. The schematic at the top shows the domain architecture of Nsp12. (Left) Ribbon representation of the crystal structure of Nsp12-remdesivir monophosphate-RNA complex (PDB code: 7BV2). The structure in the right panel shows a magnified view of the boxed area in the left panel. The Nsp12 domains are colored as in the schematic at the top. The catalytic site in Nsp12 is marked by a black circle in the right panel. The side chains of amino acids comprising the catalytic site of RdRp (Ser758, Asp759, and Asp760) are shown as balls and stick and colored yellow. The nucleotide binding site is boxed in the right panel. The side chains of amino acids participating in nucleotide binding (Lys544, Arg552, and Arg554) are shown as balls and sticks and colored light blue. Remdesivir molecule incorporated into the nascent RNA is shown as balls and sticks and colored light pink. The RNA is shown as a blue cartoon and bases are shown as sticks. The positions of Cα atoms of amino acids identified in this study are shown as red and green spheres and labeled. The amino acids that are shown as red spheres are located above the nucleotide binding site, whereas Cys812 located at the catalytic site is shown as a green sphere. The side chain of active site residue Ser758 is shown as ball and sticks and colored yellow. The location of Cα atoms of remdesivir resistance conferring amino acid Val556 is shown as blue sphere and labeled.

Importantly, none of the observed amino acid polymorphisms in RdRp were located precisely at two sites known to cause *in vitro* resistance to remdesivir (26). Most of the amino acid changes are located distantly from the RNA-binding and catalytic sites (**Figure S4** and **Table 1**). However, replacements at six amino acid residues (Ala442Val, Ala448Val, Ala553Pro/Val, Gly682Arg, Ser758Pro, and Cys812Phe) may potentially interfere with either remdesivir binding or RNA synthesis. Four (Ala442Val, Ala448Val, Ala553Pro/Val, and Gly682Arg) of the six substitution sites are located immediately above the nucleotide-binding site, that is comprised of Lys544, Arg552, and Arg554 residues as shown by structural studies (**Figure 4**). The positions of these four variant amino acid sites are comparable to Val556 (**Figure 4**), for which a Val556Leu mutation in SARS-CoV was identified to confer resistance to remdesivir *in vitro* (26). The other two substitutions (Ser758Pro and Cys812Phe) are inferred to be located either at, or in the immediate proximity of, the catalytic active site, that is comprised of three contiguous residues (Ser758, Asp759, and Asp760). A proline substitution we identified at Ser758 (Ser758Pro) is likely to negatively impact RNA synthesis. Although Cys812 is not directly involved in the catalysis of RNA synthesis, it is only 3.5 Å away from Asp760.The introduction of the bulkier phenylalanine substitution at Cys812 (Cys812Phe) may impair RNA synthesis. Consequently, these two substitutions are expected to detrimentally affect virus replication or fitness.

### Analysis of the gene encoding the spike protein

The densely glycosylated spike protein of SARS-CoV-2 and its close coronavirus relatives binds directly to host-cell angiotensin-converting enzyme 2 (ACE2) receptors to enter host cells (35-37). Thus, the spike protein is a major translational research target, including intensive vaccine and therapeutic antibody (35-64). Analysis of the gene encoding the spike protein identified 470 SNPs, including 285 that produce amino acid changes (**Table 2, Figure 5**). Forty-nine of these replacements (V11A, T51A, W64C, I119T, E156Q, S205A, D228G, L229W, P230T, N234D, I235T, T274A, A288V, E324Q, E324V, S325P, S349F, S371P, S373P, T385I, A419V, C480F, Y495S, L517F, K528R, Q628E, T632I, S708P, T719I, P728L, S746P, E748K, G757V, V772A, K814R, D843N, S884A, M902I, I909V, E918Q, S982L, M1029I, Q1142K, K1157M, Q1180R, D1199A, C1241F, C1247G, and V1268A) are not represented in a publicly available database (34) as of August 19, 2020. Interestingly, 25 amino acid sites have three distinct variants (that is, the reference amino acid plus two additional variant amino acids), and five amino acid sites (amino acid positions 21, 27, 228, 936, and 1050) have four distinct variants represented in our sample of 5,085 genomes (**Table 2, Figure 5**).

**Table 2.**
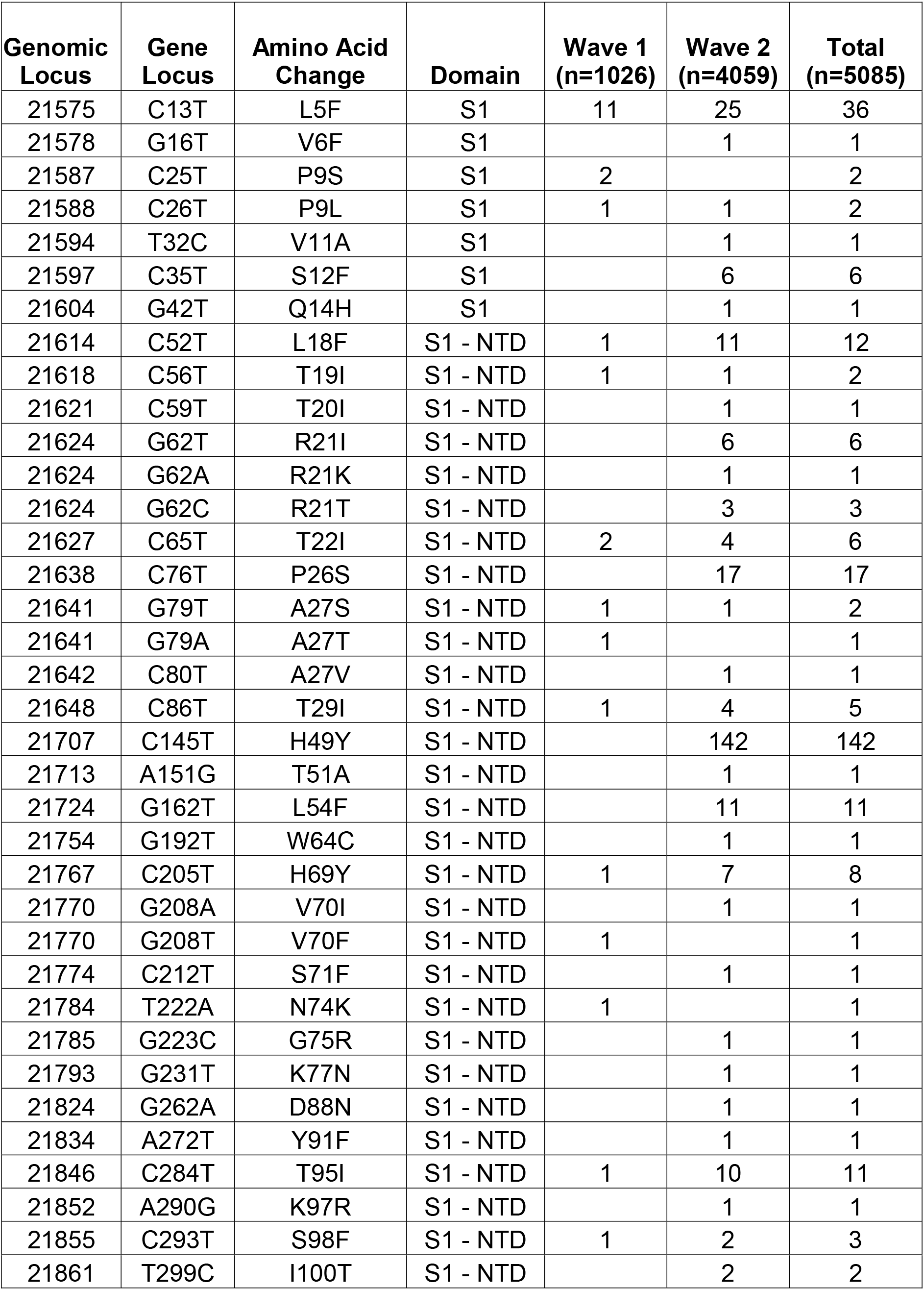
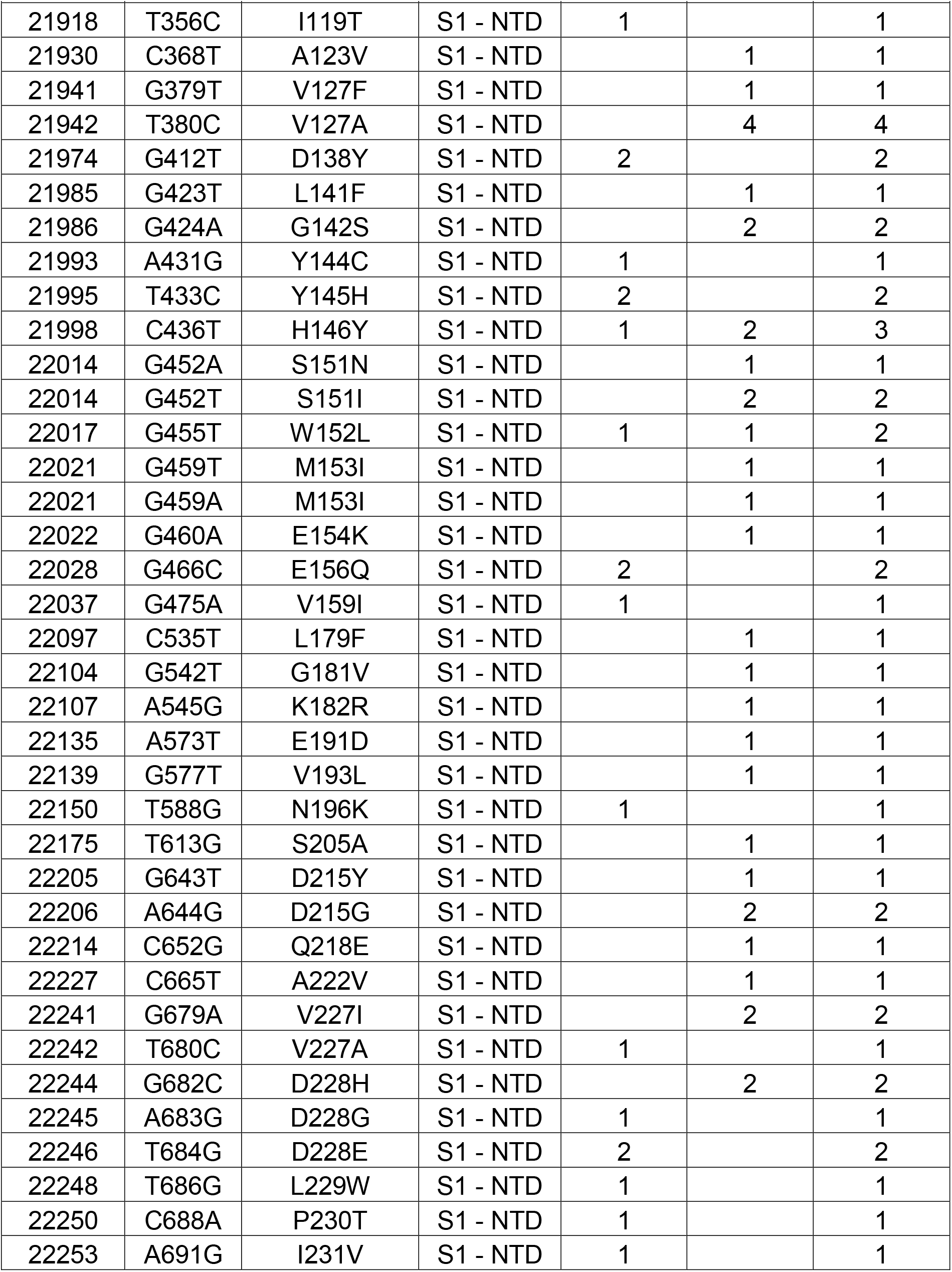
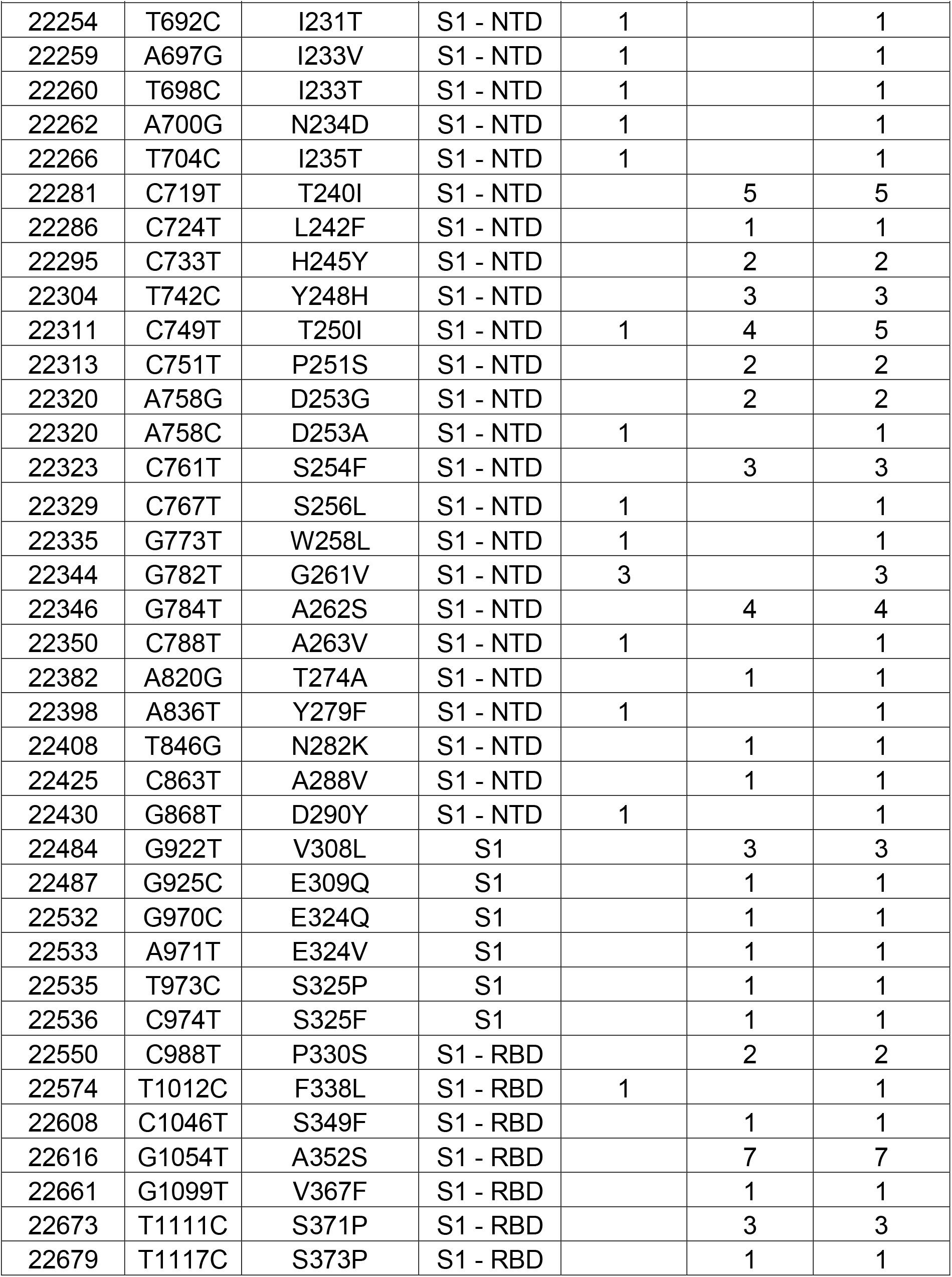
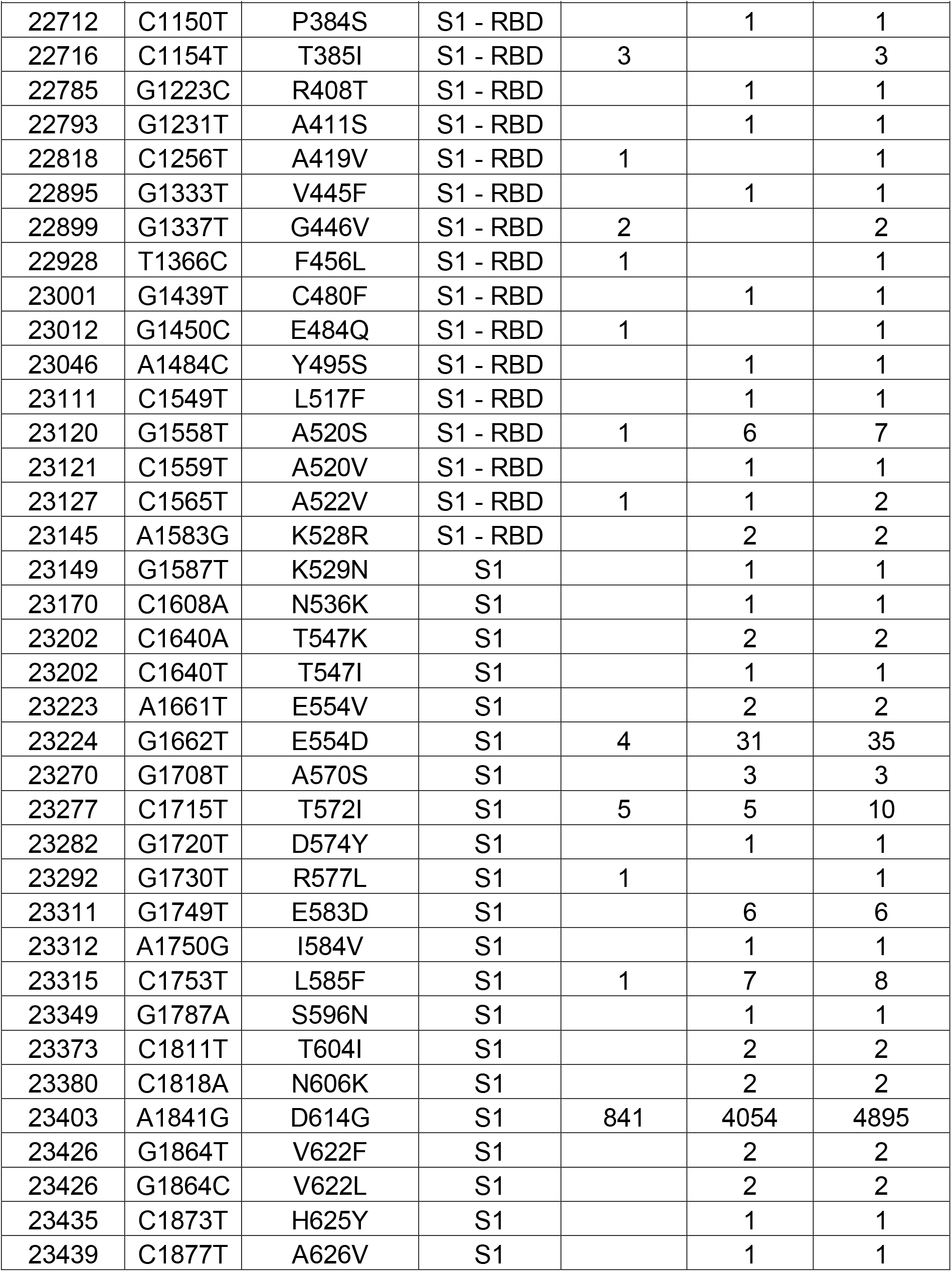
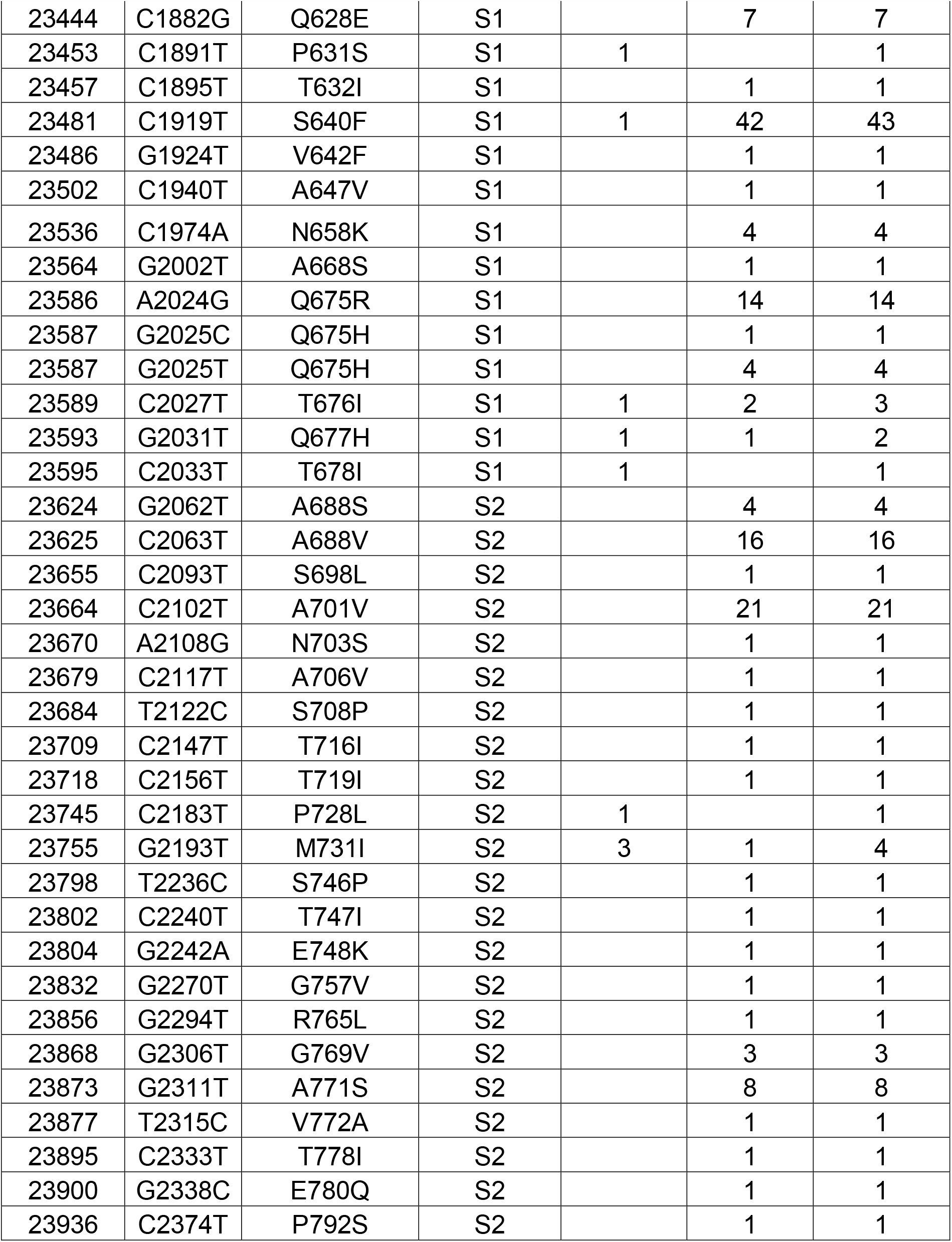
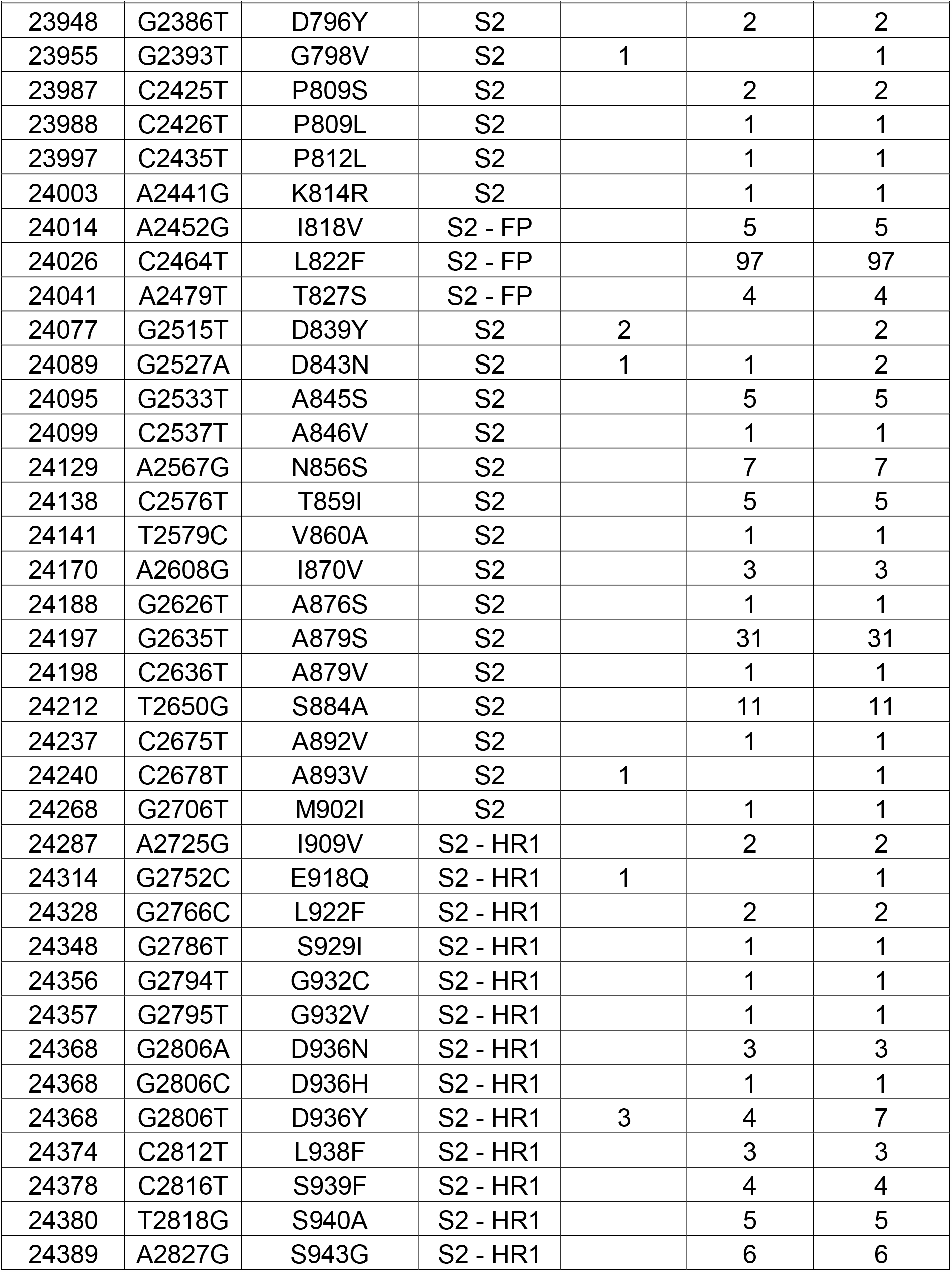
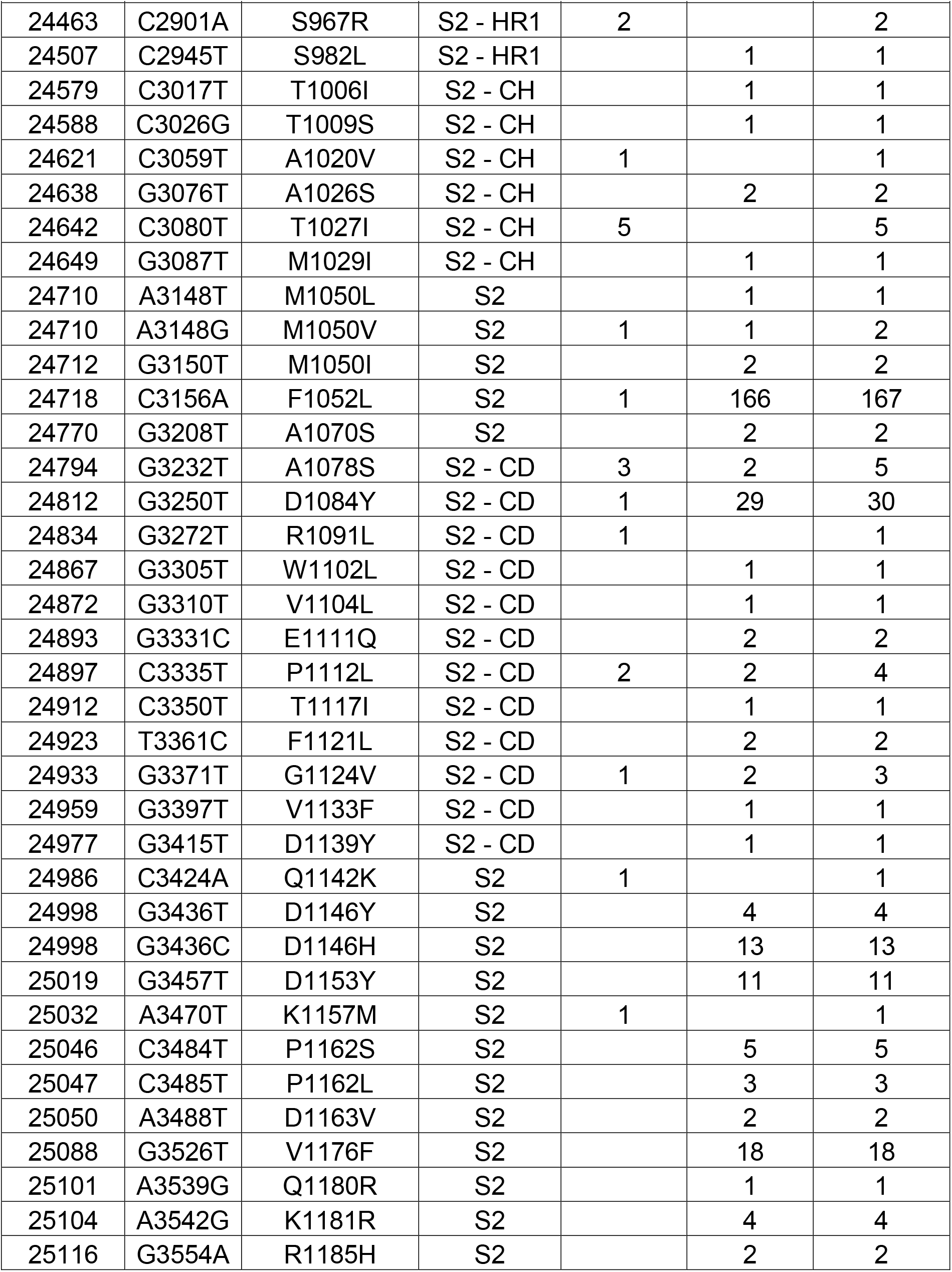
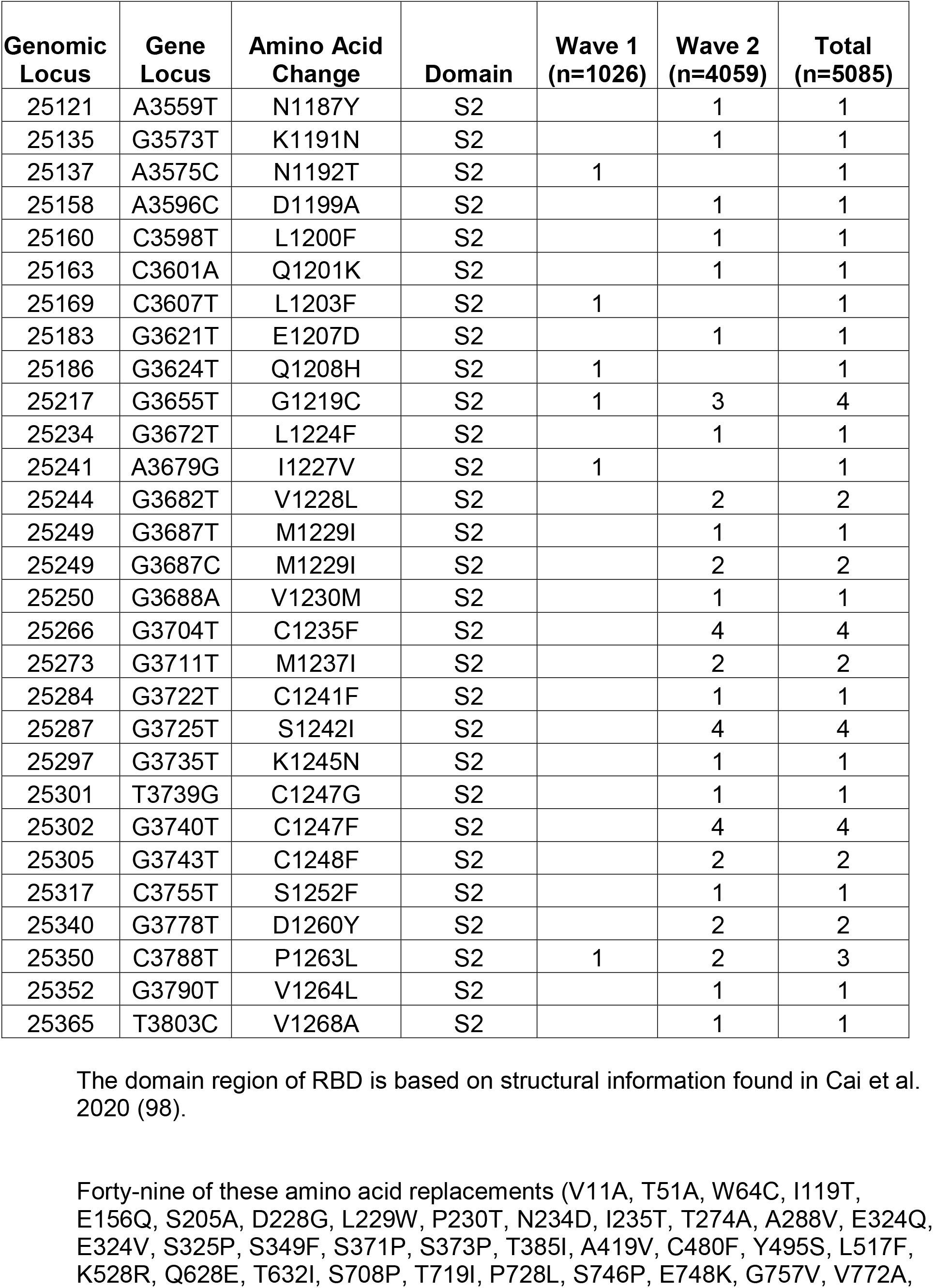
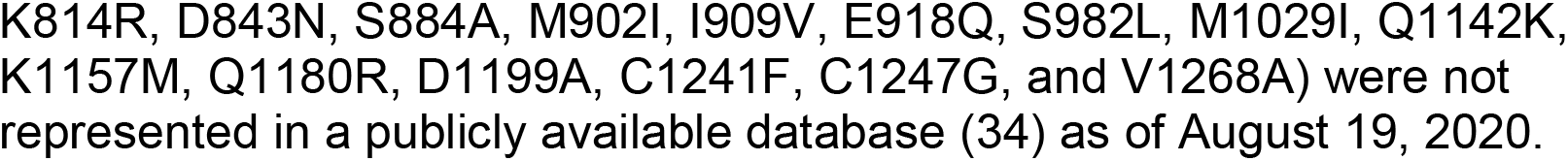
Nonsynonymous SNPs in SARS-CoV-2 spike protein

**FIG 5.**
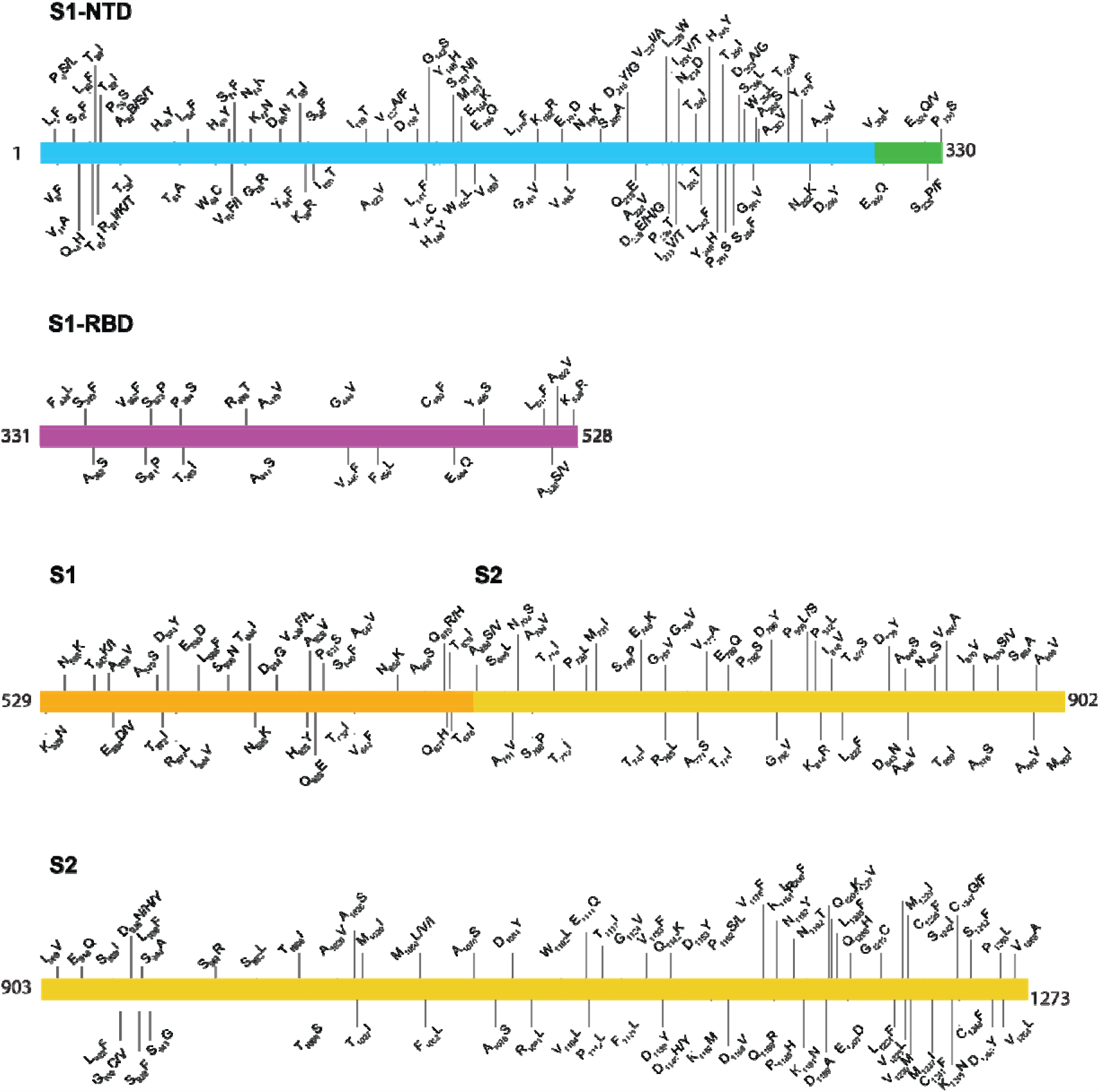
Location of amino acid replacements in spike protein among the 5,085 genomes of SARS-CoV-2 sequenced. The various spike protein domains are color-coded. The numbers refer to amino acid site. Note that many amino acid sites have multiple variants identified.

We mapped the location of amino acid replacements onto a model of the full-length spike protein (35, 65) and observed that the substitutions are found in each subunit and domain of the spike (**Figure 6**). However, the distribution of amino acid changes is not uniform throughout the protein regions. For example, compared to some other regions of the spike protein, the RBD has relatively few amino acid changes, and the frequency of strains with these substitutions is low, each occurring in fewer than 10 isolates. This finding is consistent with the functional constraints on RBD to mediate interaction with ACE2. In contrast, the periphery of the S1 subunit NTD contains a dense cluster of substituted residues, with some single amino acid replacements found in 10–20 isolates (**Table 2, Figure 5, Figure 6**). Clustering of amino acid changes in a distinct region of the spike protein may be a signal of positive selection. Inasmuch as infected patients make antibodies against the NTD, we favor the idea that host immune selection is one force contributing to some of the amino acid variation in this region. One NTD substitution, H49Y, was found in 142 isolates. This position is not well exposed on the surface of the NTD and is likely not a result of immune pressure. The same is true for another highly represented substitution, F1052L. This substitution was observed in 167 isolates, and F1052 is buried within the core of the S2 subunit. The substitution observed most frequently in the spike protein in our sample is D614G, a change observed in 4,895 of the isolates. As noted above, strains with the Gly614 variant significantly increased in wave 2 compared to wave 1.

**FIG 6.**
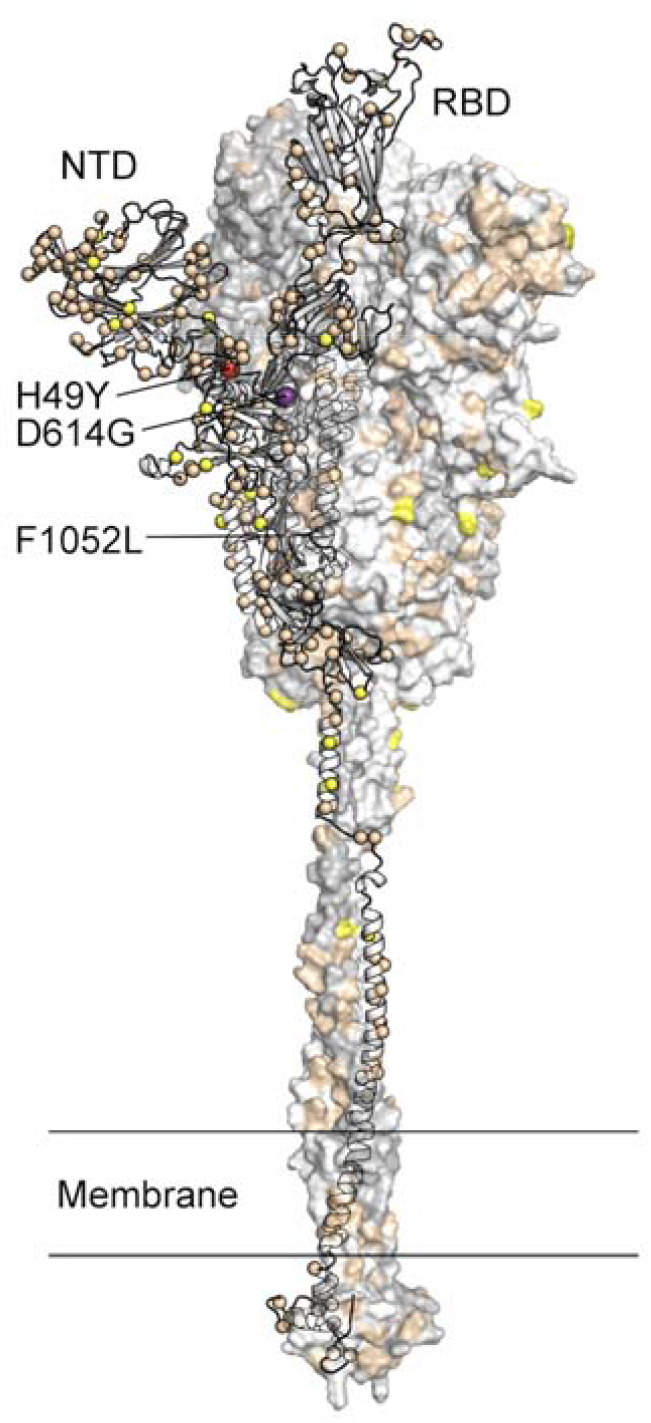
Location of amino acid substitutions mapped on the SARS-CoV-2 spike protein. Model of the SARS-CoV-2 spike protein with one protomer shown as ribbons and the other two protomers shown as a molecular surface. The Cα atom of residues found to be substituted in one or more virus isolates identified in this study is shown as a sphere on the ribbon representation. Residues found to be substituted in 1–9 isolates are colored tan, 10–99 isolates yellow, 100–999 isolates colored red (H49Y and F1052L), and >1000 isolates purple (D614G). The surface of the aminoterminal domain (NTD) that is distal to the trimeric axis has a high density of substituted residues. RBD, receptor binding domain.

As observed with RdRp, the majority of strains with each single amino acid change in the spike protein were found on a distinct phylogenetic lineage (**Figure S5**), indicating identity by descent. A prominent exception is the Leu5Phe replacement that is present in all major clades, suggesting that this amino acid change arose multiple times independently or very early in the course of SARS-CoV-2 evolution. Finally, we note that examination of the phylogenetic distribution of strains with multiple distinct amino acid replacements at the same site (e.g., Arg21Ile/Lys/Thr, Ala27Ser/Thr/Val, etc.) revealed that they were commonly found in different genetic branches, consistent with independent origin (**Figure S5**).

### Cycle threshold (Ct) comparison of SARS-CoV-2 strains with either the Asp614 or Gly614 amino acid replacements in spike protein

It has been reported that patients infected with strains having spike protein Gly614 variant have, on average, higher virus loads on initial diagnosis (66-70). To determine if this is the case in Houston strains, we examined the cycle threshold (Ct) for every sequenced strain that was detected from a patient specimen using the SARS-CoV-2 Assay done by the Hologic Panther instrument. We identified a significant difference (*p*<0.0001) between the mean Ct value for strains with an Asp614 (*n*=102) or Gly614 (*n*=812) variant of the spike protein (**Figure 7**). Strains with Gly614 had a Ct value significantly lower than strains with the Asp614 variant, indicating that patients infected with the Gly614 strains had, on average, higher virus loads on initial diagnosis than patients infected by strains with the Asp614 variant (**Figure 7**). This observation is consistent with the conjecture that, on average, strains with the Gly614 variant are better able to disseminate.

**FIG 7.**
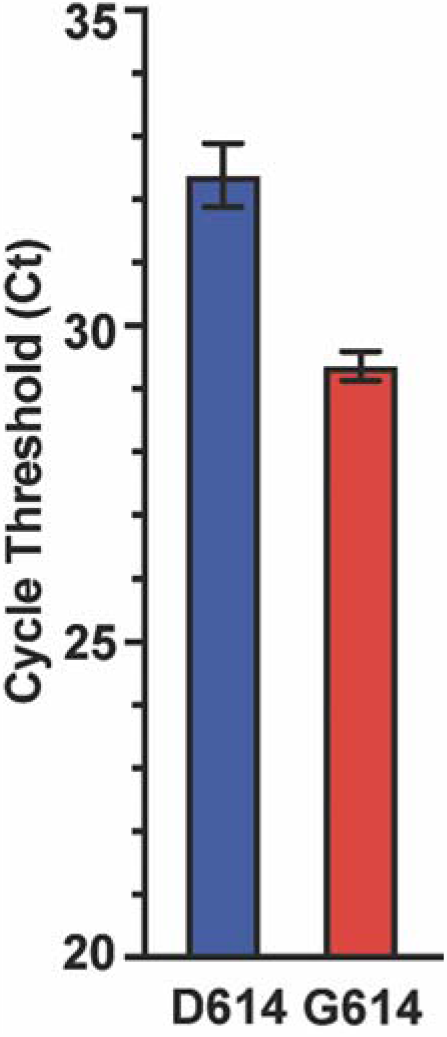
Cycle threshold (Ct) for every SARS-CoV-2 patient sample tested using the Hologic Panther assay. Data are presented as mean +/- standard error of the mean for strains with an aspartate (D614, *n*=102 strains, blue) or glycine (G614, *n*=812 strains, red) at amino acid 614 of the spike protein. Mann-Whitney test, ^*^*P*<0.0001.

### Characterization of recombinant proteins with single amino acid replacements in the receptor binding domain region of spike protein

The RBD of spike protein binds the ACE2 surface receptor and is also targeted by neutralizing (36, 37, 41, 43-46, 48-62, 71). Thus, single amino acid replacements in this domain may have functional consequences that enhance virus fitness. To begin to test this idea, we expressed spike variants with the Asp614Gly replacement and 13 clinical RBD variants identified in our genome sequencing studies (**Figure 8, Table S4A, B**). All RBD variants were cloned into an engineered spike protein construct that stabilizes the perfusion state and increases overall expression yield (spike-6P, here referred to as spike) (64).

**FIG 8.**
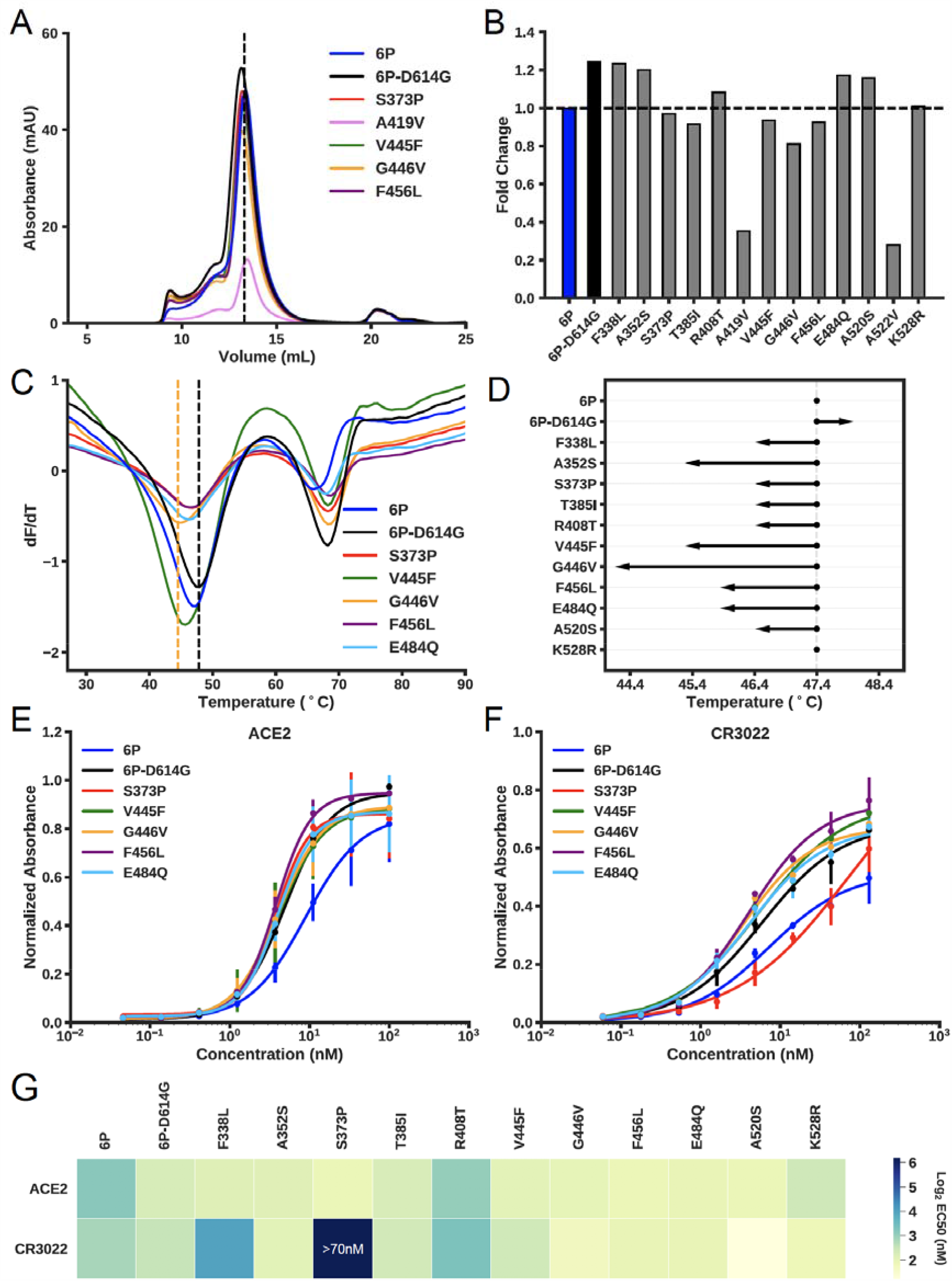
Biochemical characterization of spike RBD variants. (A) Size-exclusion chromatography (SEC) traces of the indicated spike-RBD variants. Dashed line indicates the elution peak of spike-6P. (B) The relative expression of all RBD variants as determined by the area under the SEC traces. All expression levels are normalized relative to spike-6P. (C) Thermostability analysis of RBD variants by differential scanning fluorimetry. Each sample had three replicates and only mean values were plotted. Black vertical dashed line indicates the first melting temperature of 6P-D614G and orange vertical dashed line indicates the first melting temperature of the least stable variant (spike-G446V). (D) First apparent melting temperature of all RBD variants. (E) ELISA-based binding affinities for ACE2 and (F) the neutralizing antibody CR3022 to the indicated RBD variants.(G) Summary of EC50s for all measured RBD variants.

We first assessed the biophysical properties of spike-Asp614Gly, an amino acid polymorphism that is common globally and increased significantly in our wave 2 strain isolates. Pseudotyped viruses expressing spike-Gly614 have higher infectivity for host cells *in vitro* than spike-Asp614 (66, 67, 69, 72, 73). The higher infectivity of spike-Gly614 is correlated with increased stability and incorporation of the spike protein into the pseudovirion (73). We observed a higher expression level (**Figure 8A, B**) and increased thermostability for the spike protein construct containing this variant (**Figure 8C, D**). The size exclusion chromatography (SEC) elution profile of spike-Asp614 was indistinguishable from spike-Gly614, consistent with a trimeric conformation (**Figure 8A**). These results are broadly consistent with higher-resolution structural analyses of both spike variants.

Next, we purified and biophysically characterized 13 RBD mutants that each contain Gly614 and one additional single amino acid replacement we identified by genome sequencing our clinical samples (**Table S4C**). All variants eluted as trimers, indicating the global structure, remained intact (**Figure 8** and **Figure S6**). However, several variants had reduced expression levels and virtually all had decreased thermostability relative to the variant that had only a D614G single amino acid replacement (**Figure 8D**). The A419V and A522V mutations were especially deleterious, reducing yield and precluding further downstream analysis (**Figure 8B**). We next assayed the affinity of the 11 highest-expressing spike variants for ACE2 and the neutralizing monoclonal antibody CR3022 via enzyme-linked immunosorbent assays (ELISAs) (**Figure 8E-G** and **Table S4C**). Most variants retained high affinity for the ACE2 surface receptor. However, importantly, three RBD variants (F338L, S373P, and R408T) had substantially reduced affinity for CR3022, a monoclonal antibody that disrupts the spike protein homotrimerization interface (63, 74). Notably, the S373P mutation is one amino acid away from the epitope recognized by CR3022. These results are consistent with the interpretation that some RBD mutants arising in COVID-19 patients may have increased ability to escape humoral immune pressure, but otherwise retain strong ACE2 binding affinity.

## DISCUSSION

In this work we analyzed the molecular population genomics, sociodemographic, and medical features of two waves of COVID-19 disease occurring in metropolitan Houston, Texas, between early March and early July 2020. We also studied the biophysical and immunologic properties of some naturally occurring single amino acid changes in the spike protein RBD identified by sequencing the 5,085 genomes. We discovered that the first COVID-19 wave was caused by a heterogenous array of virus genotypes assigned to several different clades. The majority of cases in the first wave are related to strains that caused widespread disease in European and Asian countries, as well as other localities. We conclude that the SARS-CoV-2 virus was introduced into Houston many times independently, likely by individuals who had traveled to or from different parts of the world, including other communities in the United States. In support of this conclusion, the first cases in metropolitan Houston were associated with a travel history to a known COVID-19 region (16). The data are consistent with the fact that Houston is a large international city characterized by a multi-ethnic population and is a prominent transport hub with direct flights to major cities globally.

The second wave of COVID-19 cases also is characterized by SARS-CoV-2 strains with diverse genotypes. Virtually all cases in the second and ongoing disease wave were caused by strains with the Gly614 variant of spike protein (**Figure 1B**). Our data unambiguously demonstrate that strains with the Gly614 variant increased significantly in frequency in wave 2 relative to wave 1 in the Houston metropolitan region. This shift occurred very rapidly in a matter of just a few months. Amino acid residue Asp614 is located in subdomain 2 (SD-2) of the spike protein and forms a hydrogen bond and electrostatic interaction with two residues in the S2 subunit of a neighboring protomer. Replacement of aspartate with glycine would eliminate both interactions, thereby substantively weakening the contact between the S1 and S2 subunits. We previously speculated (75) that this weakening produces a more fusogenic spike protein, as S1 must first dissociate from S2 before S2 can refold and mediate fusion of virus and cell membranes. Stated another way, virus strains with the Gly614 variant may be better able to enter host cells, potentially resulting in enhanced spread.Consistent with this idea, Korber et al. (66) showed that the Gly614 variant grows to higher titer as pseudotyped virions. On initial diagnosis infected individuals had lower RT-PCR cycle thresholds suggesting higher upper respiratory tract viral loads. Our data (**Figure 7**) are fully consistent with that finding Zhang et al. (73) reported that pseudovirus with the 614Gly variant infected ACE2-expressing cells more efficiently than the 614Asp. Similar results have been described by Hu et al. (67) and Lorenzo-Redondo et al. (68). Plante et al. (76) recently studied isogenic mutant SARS-CoV-2 strains with either the 614Asp or 614Gly variant and found that the 614Gly variant virus had significantly increased replication in human lung epithelial cells *in vitro* and increased infectious titers in nasal and trachea washes obtained from experimentally infected hamsters. These results are consistent with the idea that the 614Gly variant bestows increased virus fitness in the upper respiratory tract (76).

Additional work is needed to investigate the potential biomedical relevance and public health importance of the Asp614Gly polymorphism, including but not limited to virus dissemination, overall fitness, impact on clinical course and virulence, and development of vaccines and therapeutics. Although it is possible that stochastic processes alone may account for the rapid increase in COVID-19 disease frequency caused by viruses containing the Gly614 variant, we do not favor that interpretation in part because of the cumulative weight of the epidemiologic, human RT-PCR diagnostics data, *in vitro* experimental findings, and animal infection studies using isogenic mutant virus strains. In addition, if stochastic processes solely are responsible, we believe it is difficult to explain essentially simultaneous increase in frequency of the Gly614 variant in genetically diverse viruses in three distinct clades (G, GH, and GR) in a geographically large metropolitan area with 7 million ethnically diverse people. Regardless, more research on this important topic is warranted.

The diversity present in our 1,026 virus genomes from the first disease wave contrasts somewhat with data reported by Gonzalez-Reiche et al., who studied 84 SARS-CoV-2 isolates causing disease in patients in the New York City region (11). Those investigators concluded that the vast majority of disease was caused by progeny of strains imported from Europe. Similarly, Bedford et al.(10) reported that much of the COVID-19 disease in the Seattle, Washington area was caused by strains that are progeny of a virus strain recently introduced from China. Some aspects of our findings are similar to those reported recently by Lemieux et al. based on analysis of strains causing disease in the Boston area (81). Our findings, like theirs, highlight the importance of multiple importation events of genetically diverse strains in the epidemiology of COVID-19 disease in this pandemic. Similarly, Icelandic and Brazilian investigators documented that SARS-CoV-2 was imported by individuals traveling to or from many European and other countries (82, 83).

The virus genome diversity and large sample size in our study permitted us to test the hypothesis that distinct virus clades were nonrandomly associated with hospitalized COVID-19 patients or disease severity. We did not find evidence to support this hypothesis, but our continuing study of COVID-19 cases accruing in the second wave will further improve statistical stratification.

We used machine learning classifiers to identify if any SNPs contribute to increased infection severity or otherwise affect virus-host outcome. The models could not be trained to accurately predict these outcomes from the available virus genome sequence data. This may be due to sample size or class imbalance.

However, we do not favor this interpretation. Rather, we think that the inability to identify particular virus SNPs predictive of disease severity or infection outcome likely reflects the substantial heterogeneity in underlying medical conditions and treatment regimens among COVID-19 patients studied herein. An alternative but not mutually exclusive hypothesis is that patient genotypes play an important role in determining virus-human interactions and resulting pathology. Although some evidence has been presented in support of this idea (84, 85), available data suggest that in the aggregate, host genetics does not play an overwhelming role in determining outcome in the great majority of adult patients, once virus infection is established.

Remdesivir is a nucleoside analog reported to have activity against MERS-CoV, a coronavirus related to SARS-CoV-2. Recently, several studies have reported that remdesivir shows promise in treating COVID-19 patients (29-33), leading the FDA to issue an emergency use authorization. Because *in vitro* resistance of SARS-CoV to remdesivir has been reported to be caused by either of two amino acid replacements in RdRp (Phe479Leu or Val556Leu), we interrogated our data for polymorphisms in the *nsp12* gene. Although we identified 140 different inferred amino acid replacements in RdRp in the 5,085 genomes analyzed, none of these were located precisely at the two positions associated with *in vitro* resistance to remdesivir. Inasmuch as remdesivir is now being deployed widely to treat COVID-19 patients in Houston and elsewhere, our findings suggest that the majority of SARS-CoV-2 strains currently circulating in our region should be susceptible to this drug.

The amino acid replacements Ala442Val, Ala448Val, Ala553Pro/Val, and Gly682Arg that we identified occur at sites that, intriguingly, are located directly above the nucleotide substrate entry channel and nucleotide binding residues Lys544, Arg552, and Arg554 (22, 23) (**Figure 4**). One possibility is that substitution of the smaller alanine or glycine residues with the bulkier side chains of Val/Pro/Arg may impose structural constraints for the modified nucleotide analog to bind, and thereby disfavor remdesivir binding. This, in turn, may lead to reduced incorporation of remdesivir into the nascent RNA, increased fidelity of RNA synthesis, and ultimately drug resistance. A similar mechanism has been proposed for a Val556Leu change (23).

We also identified one strain with a Lys477Asn replacement in RdRp. This substitution is located close to a Phe479Leu replacement reported to produce partial resistance to remdesivir *in vitro* in SARS-CoV patients from 2004, although the amino acid positions are numbered differently in SARS-CoV and SARS-CoV-2. Structural studies have suggested that this amino acid is surface-exposed, and distant from known key functional elements. Our observed Lys477Asn change is also located in a conserved motif described as a finger domain of RdRp (**Figure 3** and **4**). One speculative possibility is that Lys477 is involved in binding a yet unidentified cofactor such as Nsp7 or Nsp8, an interaction that could modify nucleotide binding and/or fidelity at a distance. These data warrant additional study in larger patient cohorts, especially in individuals treated with remdesivir.

Analysis of the gene encoding the spike protein identified 285 polymorphic amino acid sites relative to the reference genome, including 49 inferred amino acid replacements not present in available databases as of August 19, 2020. Importantly, 30 amino acid sites in the spike protein had two or three distinct replacements relative to the reference strain. The occurrence of multiple variants at the same amino acid site is one characteristic that may suggest functional consequences. These data, coupled with structural information available for spike protein, raise the possibility that some of the amino acid variants have functional consequences, for example including altered serologic reactivity and shown here. These data permit generation of many biomedically relevant hypotheses now under study.

A recent study reported that RBD amino acid changes could be selected *in vitro* using a pseudovirus neutralization assay and sera obtained from convalescent plasma or monoclonal antibodies (86). The amino acid sites included positions V445 and E484 in the RBD. Important to note, variants G446V and E484Q were present in our patient samples. However, these mutations retain high affinity to CR3022 **(Figure 8F, G**). The high-resolution structure of the RBD/CR3022 complex shows that CR3022 makes contacts to residues 369-386, 380-392, and 427-430 of RBD (74). Although there is no overlap between CR3022 and ACE2 epitopes, CR3022 is able to neutralize the virus through an allosteric effect. We found that the Ser373Pro change, which is located within the CR3022 epitope, has reduced affinity to CR3022 (**Figure 8F, G**). The F338L and R408T mutations, although not found directly within the interacting epitope, also display reduced binding to CR3022. Other investigators (86) using *in vitro* antibody selection identified a change at amino acid site S151 in the N-terminal domain, and we found mutations S151N and S151I in our patient samples. We also note that two variant amino acids (Gly446Val and Phe456Leu) we identified are located in a linear epitope found to be critical for a neutralizing monoclonal antibody described recently by Li et al. (87).

In the aggregate, these findings suggest that mutations emerging within the spike protein at positions within and proximal to known neutralization epitopes may result in escape from antibodies and other therapeutics currently under development. Importantly, our study did not reveal that these mutant strains had disproportionately increased over time. The findings may also bear on the occurrence of multiple amino acid substitutions at the same amino acid site that we identified in this study, commonly a signal of selection. In the aggregate, the data support a multifaceted approach to serological monitoring and biologics development, including the use of monoclonal antibody cocktails (46, 47, 88).

## CONCLUDING STATEMENT

Our work represents analysis of the largest sample to date of SARS-CoV-2 genome sequences from patients in one metropolitan region in the United States. The investigation was facilitated by the fact that we had rapidly assessed a SARS-CoV-2 molecular diagnostic test in January 2020, more than a month before the first COVID-19 patient was diagnosed in Houston. In addition, our large healthcare system has seven hospitals and many facilities (e.g., outpatient care centers, emergency departments) located in geographically diverse areas of the city. We also provide reference laboratory services for other healthcare entities in the Houston area. Together, our facilities serve patients of diverse ethnicities and socioeconomic status. Thus, the data presented here likely reflect a broad overview of virus diversity causing COVID-19 infections throughout metropolitan Houston. We previously exploited these features to study influenza and *Klebsiella pneumoniae* dissemination in metropolitan Houston (89, 90). We acknowledge that every “twig” of the SARS-CoV-2 evolutionary tree in Houston is not represented in these data. The samples studied are not comprehensive for the entire metropolitan region. For example, it is possible that our strain samples are not fully representative of individuals who are indigent, homeless, or of very low socioeconomic groups. In addition, although the strain sample size is relatively large compared to other studies, the sample represents only about 10% of all COVID-19 cases in metropolitan Houston documented in the study period. In addition, some patient samples contain relatively small amounts of virus nucleic acid and do not yield adequate sequence data for high-quality genome analysis. Thus, our data likely underestimate the extent of genome diversity present among SARS-CoV-2 causing COVID-19 and will not identify all amino acid replacements in the virus in this geographic region. It will be important to sequence and analyze the genomes of additional SARS-CoV-2 strains causing COVID-19 cases in the ongoing second massive disease wave in metropolitan Houston, and these studies are underway. Data of this type will be especially important to have if a third and subsequent waves were to occur in metropolitan Houston, as it could provide insight into molecular and epidemiologic events contributing to them.

The genomes reported here are an important data resource that will underpin our ongoing study of SARS-CoV-2 molecular evolution, dissemination, and medical features of COVID-19 in Houston. As of August 19, 2020, there were 135,866 reported cases of COVID-19 in metropolitan Houston, and the number of cases is increasing daily. Although the full array of factors contributing to the massive second wave in Houston is not known, it is possible that the potential for increased transmissibility of SARS-CoV-2 with the Gly614 may have played a role, as well as changes in behavior associated with the Memorial Day and July 4^th^ holidays, and relaxation of some of the social constraints imposed during the first wave. The availability of extensive virus genome data dating from the earliest reported cases of COVID-19 in metropolitan Houston, coupled with the database we have now constructed, may provide critical insights into the origin of new infection spikes and waves occurring as public health constraints are further relaxed, schools and colleges re-open, holidays occur, commercial air travel increases, and individuals change their behavior because of COVID-19 “fatigue.” The genome data will also be useful in assessing ongoing molecular evolution in spike and other proteins as baseline herd immunity is generated, either by natural exposure to SARS-CoV-2 or by vaccination. The signal of potential selection contributing to some spike protein diversity and identification of naturally occurring mutant RBD variants with altered serologic recognition warrant close attention and expanded study.

## MATERIALS AND METHODS

### Patient specimens

All specimens were obtained from individuals who were registered patients at Houston Methodist hospitals, associated facilities (e.g., urgent care centers), or institutions in the greater Houston metropolitan region that use our laboratory services. Virtually all individuals met the criteria specified by the Centers for Disease Control and Prevention to be classified as a person under investigation.

### SARS-CoV-2 molecular diagnostic testing

Specimens obtained from symptomatic patients with a high degree of suspicion for COVID-19 disease were tested in the Molecular Diagnostics Laboratory at Houston Methodist Hospital using an assay granted Emergency Use Authorization (EUA) from the FDA (https://www.fda.gov/medical-devices/emergency-situations-medical-devices/faqs-diagnostic-testing-sars-cov-2#offeringtests). Multiple testing platforms were used, including an assay that follows the protocol published by the WHO (https://www.who.int/docs/default-source/coronaviruse/protocol-v2-1.pdf) using the EZ1 virus extraction kit and EZ1 Advanced XL instrument or QIASymphony DSP Virus kit and QIASymphony instrument for nucleic acid extraction and ABI 7500 Fast Dx instrument with 7500 SDS software for reverse transcription RT-PCR, the COVID-19 test using BioFire Film Array 2.0 instruments, the Xpert Xpress SARS-CoV-2 test using Cepheid GeneXpert Infinity or Cepheid GeneXpert Xpress IV instruments, the SARS-CoV-2 Assay using the Hologic Panther instrument, and the Aptima SARS-CoV-2 Assay using the Hologic Panther Fusion system. All assays were performed according to the manufacturer’s instructions. Testing was performed on material obtained from nasopharyngeal or oropharyngeal swabs immersed in universal transport media (UTM), bronchoalveolar lavage fluid, or sputum treated with dithiothreitol (DTT). To standardize specimen collection, an instructional video was created for Houston Methodist healthcare workers (https://vimeo.com/396996468/2228335d56).

### Epidemiologic curve

The number of confirmed COVID-19 positive cases was obtained from USAFacts.org (https://usafacts.org/visualizations/coronavirus-covid-19-spread-map/) for Austin, Brazoria, Chambers, Fort Bend, Galveston, Harris, Liberty, Montgomery, and Waller counties. Positive cases for Houston Methodist Hospital patients were obtained from our Laboratory Information System and plotted using the documented collection time.

### SARS-CoV-2 genome sequencing

Libraries for whole virus genome sequencing were prepared according to version 1 or version 3 of the ARTIC nCoV-2019 sequencing protocol (https://artic.network/ncov-2019). Long reads were generated with the LSK-109 sequencing kit, 24 native barcodes (NBD104 and NBD114 kits), and a GridION instrument (Oxford Nanopore). Short reads were generated with the NexteraXT kit and NextSeq 550 instrument (Illumina).

### SARS-CoV-2 genome sequence analysis

Consensus virus genome sequences from the Houston area isolates were generated using the ARTIC nCoV-2019 bioinformatics pipeline. Publicly available genomes and metadata were acquired through GISAID on August 19, 2020. GISAID sequences containing greater than 1% N characters, and Houston sequences with greater than 5% N characters were removed from consideration. Identical GISAID sequences originating from the same geographic location with the same collection date were also removed from consideration to reduce redundancy. Nucleotide sequence alignments for the combined Houston and GISAID strains were generated using MAFFT version 7.130b with default parameters (91). Sequences were manually curated in JalView (92) to trim the ends and to remove sequences containing spurious inserts. Phylogenetic trees were generated using FastTree with the generalized time-reversible model for nucleotide sequences (93). CLC Genomics Workbench (QIAGEN) was used to generate the phylogenetic tree figures.

### Geospatial mapping

The home address zip code for all SARS-CoV-2 positive patients was used to generate the geospatial maps. To examine geographic relatedness among genetically similar isolates, geospatial maps were filtered to isolates containing specific amino acid changes.

### Time series

Geospatial data were filtered into wave 1 (3/5/2020-5/11/2020) and wave 2 (5/12/2020-7/7/2020) time intervals to illustrate the spread of confirmed SARS-CoV-2 positive patients identified over time.

### Machine learning

Virus genome alignments and patient metadata were used to build models to predict patient metadata and outcomes using both classification models and regression. Metadata considered for prediction in the classification models included age, ABO and Rh blood type, ethnic group, ethnicity, sex, ICU admission, IMU admission, supplemental oxygen use, and ventilator use. Metadata considered for prediction in regression analysis included ICU length of stay, IMU length of stay, total length of stay, supplemental oxygen use, and ventilator use. Because sex, blood type, Rh factor, age, age decade, ethnicity, and ethnic group are features in the patient features and combined feature sets, models were not trained for these labels using patient and combined feature sets. Additionally, age, length of stay, IMU length of stay, ICU length of stay, mechanical ventilation days, and supplemental oxygen days were treated as regression problems and XGBoost regressors were built while the rest were treated as classification problems and XGBoost classifiers were built.

Three types of features were considered for training the XGBoost classifiers: alignment features, patient features, and the combination of alignment and patient features. Alignment features were generated from the consensus genome alignment such that columns containing ambiguous nucleotide bases were removed to ensure the models did not learn patterns from areas of low coverage. These alignments were then one-hot encoded to form the alignment features. Patient metadata values were one-hot encoded with the exception of age, which remained as a raw integer value, to create the patient features. These metadata values consisted of age, ABO, Rh blood type, ethnic group, ethnicity, and sex. All three types of feature sets were used to train models that predict ICU length of stay, IMU length of stay, overall length of stay, days of supplemental oxygen therapy, and days of ventilator usage while only alignment features were used to train models that predict age, ABO, Rh blood type, ethnic group, ethnicity, and sex.

A ten-fold cross validation was used to train XGBoost models (94) as described previously (95, 96). Depths of 4, 8, 16, 32, and 64 were used to tune the models, but accuracies plateaued after a depth of 16. SciKit-Learn’s (97) classification report and r2 score were then used to access overall accuracy of the classification and regression models, respectively.

### Patient metadata correlations

We encoded values into multiple columns for each metadata field for patients if metadata was available. For example, the ABO column was divided into four columns for A, B, AB, and O blood type. Those columns were encoded with a 1 for the patients’ ABO type, with all other columns encoded with 0. This was repeated for all non-outcome metadata fields. Age, however, was not re-encoded, as the raw integer values were used. Each column was then correlated to the various outcome values for each patient (deceased, ICU length, IMU length, length of stay, supplemental oxygen length, and ventilator length) to obtain a Pearson coefficient correlation value for each metadata label and outcome.

### Analysis of the *nsp12* polymerase and S protein genes

The *nsp12* virus polymerase and S protein genes were analyzed by plotting SNP density in the consensus alignment using Python (Python v3.4.3, Biopython Package v1.72). The frequency of SNPs in the Houston isolates was assessed, along with amino acid changes for nonsynonymous SNPs.

### Cycle threshold (Ct) comparison of SARS-CoV-2 strains with either Asp614 or Gly614 amino acid replacements in the spike protein

The cycle threshold (Ct) for every sequenced strain that was detected from a patient specimen using the SARS-CoV-2 Assay on the Hologic Panther instrument was retrieved from the Houston Methodist Hospital Laboratory Information System. Statistical significance between the mean Ct value for strains with an aspartate (*n*=102) or glycine (*n*=812) amino acid at position 614 of the spike protein was determined with the Mann-Whitney test (GraphPad PRISM 8).

### Creation and characterization of spike protein RBD variants

Spike RBD variants were cloned into the spike-6P (HexaPro; F817P, A892P, A899P, A942P, K986P, V987P) base construct that also includes the D614G substitution (pIF638). Briefly, a segment of the gene encoding the RBD was excised with EcoRI and NheI, mutagenized by PCR, and assembled with a HiFi DNA Assembly Cloning Kit (NEB).

FreeStyle 293-F cells (Thermo Fisher Scientific) were cultured and maintained in a humidified atmosphere of 37°C and 8% CO_2_ while shaking at 110-125rpm. Cells were transfected with plasmids encoding spike protein variants using polyethylenimine. Three hours post-transfection, 5μM kifunensine was added to each culture. Cells were harvested four days after transfection and the protein containing supernatant was separated from the cells by two centrifugation steps: 10 min at 500rcf and 20 min at 10,000rcf. Supernatants were kept at 4°C throughout. Clarified supernatant was loaded on a Poly-Prep chromatography column (Bio-Rad) containing Strep-Tactin Superflow resin (IBA), washed with five column volumes (CV) of wash buffer (100mM Tris-HCl pH 8.0, 150mM NaCl; 1mM EDTA), and eluted with four CV of elution buffer (100mM Tris-HCl pH 8.0, 150mM NaCl, 1mM EDTA, 2.5mM d-Desthiobiotin). The eluate was spin-concentrated (Amicon Ultra-15) to 600μL and further purified via size-exclusion chromatography (SEC) using a Superose 6 Increase 10/300 column (G.E.) in SEC buffer (2mM Tris pH 8.0, 200mM NaCl and 0.02% NaN_3_). Proteins were concentrated to 300μL and stored in SEC buffer.

The RBD spike mutants chosen for analysis were all RBD amino acid mutants identified by our genome sequencing study as of June 15, 2020. We note that the exact boundaries of the RBD domain varies depending on the paper used as reference. We used the boundaries demarcated in Figure 1A of Cai et al. Science paper 21 July) (98) that have K528R located at the RBD-CTD1 interface.

### Differential scanning fluorimetry

Recombinant spike proteins were diluted to a final concentration of 0.05mg/mL with 5X SYPRO orange (Sigma) in a 96-well qPCR plate. Continuous fluorescence measurements (λex=465nm, λem=580nm) were collected with a Roche LightCycler 480 II. The temperature was increased from 22°C to 95°C at a rate of 4.4°C/min. We report the first melting transition.

### Enzyme-linked immunosorbent assays

ELISAs were performed to characterize binding of S6P, S6P D614G, and S6P D614G-RBD variants to human ACE2 and the RBD-binding monoclonal antibody CR3022. The ACE2-hFc chimera was obtained from GenScript (Z03484), and the CR3022 antibody was purchased from Abcam (Ab273073). Corning 96-well high-binding plates (CLS9018BC) were coated with spike variants at 2μg/mL overnight at 4°C. After washing four times with phosphate buffered saline + 0.1% Tween20 (PBST; 300μL/well), plates were blocked with PBS+2% milk (PBSM) for 2 h at room temperature and again washed four times with PBST. These were serially diluted in PBSM 1:3 seven times in triplicate. After 1 h incubation at room temperature, plates were washed four times in PBST, labeled with 50μL mouse anti-human IgG1 Fc-HRP (SouthernBlots, 9054-05) for 45 min in PBSM, and washed again in PBST before addition of 50μL 1-step Ultra TMB-ELISA substrate (Thermo Scientific, 34028). Reactions were developed for 15 min and stopped by addition of 50μL 4M H_2_SO_4_. Absorbance intensity (450nm) was normalized within a plate and EC_50_ values were calculated through 4-parameter logistic curve (4PL) analysis using GraphPad PRISM 8.4.3.

## Data Availability

The SARS-CoV-2 genomes described herein have all been deposited in GISAID and are publically available.

## ACKNOWLEDGMENTS

We thank Dr. Steven Hinrichs and colleagues at the Nebraska Public Health Laboratory, and Dr. David Persse and colleagues at the Houston Health Department for providing samples used to validate our initial SARS-CoV-2 molecular assay. We thank Drs. Jessica Thomas and Zejuan Li, Erika Walker, Concepcion C. Cantu, the very talented and dedicated molecular technologists, and the many labor pool volunteers in the Molecular Diagnostics Laboratory for their dedication to patient care. We also thank Brandi Robinson, Harrold Cano, Cory Romero, Brooke Burns, and Hayder Mahmood for technical assistance. We are indebted to Drs. Marc Boom and Dirk Sostman for their support, and to many very generous Houston philanthropists for their tremendous support of this ongoing project, including but not limited to anonymous, Ann and John Bookout III, Carolyn and John Bookout, Ting Tsung and Wei Fong Chao Foundation, Ann and Leslie Doggett, Freeport LNG, the Hearst Foundations, Jerold B. Katz Foundation, C. James and Carole Walter Looke, Diane and David Modesett, the Sherman Foundation, and Paula and Joseph C. “Rusty” Walter III. We gratefully acknowledge the originating and submitting laboratories of the SARS-CoV-2 genome sequences from GISAID’s EpiFluTM Database used in some of the work presented here. We also thank many colleagues for critical reading of the manuscript and suggesting improvements, and Sasha Pejerrey, Adrienne Winston, Heather McConnell, and Kathryn Stockbauer for editorial contributions. We appreciate Dr. Stephen Schaffner for his helpful comments regarding the correlation analysis. We are especially indebted to Drs. Nancy Jenkins and Neal Copeland for their scholarly suggestions to improve an early version of the manuscript.

## Author contributions

J.M.M. conceptualized and designed the project; S.W.L, R.J.O., P.A.C., D.W.B., J.J.D., M.S., M.N., M.O.S., C.C.C., P.Y., L.P., S.S., H.-C. K., H.H., G.E., H.A.T.N., J.H.L., M.K., J.G., D.B., J.G., J.S.M., C.-W.C., K.J., and I.F. performed research. All authors contributed to writing the manuscript. Data and material availability: The spike-6P (“HexaPro”) plasmid is available from Addgene (ID: 154754) or from I.J.F. under a material transfer agreement with The University of Texas at Austin. Additional plasmids are available upon request from I.J.F.

This study was supported by the Fondren Foundation, Houston Methodist Hospital and Research Institute (to J.M.M.), NIH grant AI127521 (to J.S.M.), NIH grants GM120554 and GM124141 to I.J.F., the Welch Foundation (F-1808 to I.J.F.), and the National Science Foundation (1453358 to I.J.F.). I.J.F. is a CPRIT Scholar in Cancer Research. J.J.D., M.S., and M.N. are supported by the NIAID Bacterial and Viral Bioinformatics resource center award (contract number 75N93019C00076).

## [Supplemental Figure Legends]

**Supplemental FIG 1** Geographic distribution of representative SARS-CoV-2 subclades in the Houston metropolitan region. Blue shaded areas denote zip codes containing COVID-19 cases with the designated subclade.

**Supplemental FIG 2** Cladograms showing distribution of patient metadata, including (A) age (in decade), (B) sex, (C) ethnicity/ethnic group, (D) wave, (E) level of care, (F) mechanical ventilation, (G) length of stay, and (H) mortality.

**Supplemental FIG 3** Distribution of subclades characterized by particular amino acid replacements in Nsp12 (RdRp).

**Supplemental FIG 4** Mapping the location of amino acid replacements on Nsp12 (RdRp) from COVID-19 virus. The schematic on the top shows the domain architecture of Nsp12. The individual domains of Nsp12 are color-coded and labeled. Ribbon representation of the crystal structure of Nsp12-remdesivir monophosphate-RNA complex is shown (PDB code: 7BV2). The structure in the right panel is obtained by rotating the left panel 180° along the y-axis. The Nsp12 domains are colored as in the schematic at the top. The positions of Cα atoms of the surface-exposed amino acids identified in this study are shown as yellow spheres, whereas the positions of Cα atoms of the buried amino acids are depicted as cyan spheres. The catalytic site in RdRp is marked by a black circle in the right panel. The side chains of amino acids comprising the catalytic site of RdRp are shown as balls and sticks and colored yellow. The nucleotide binding site is boxed and labeled in the right panel. The side chains of amino acids participating in nucleotide binding (Lys545, Arg553, and Arg555) are shown as balls and sticks. Remdesivir molecule incorporated into the nascent RNA is shown as balls and sticks and colored light pink. The RNA is shown as blue cartoon and bases are shown as sticks. The positions of Cα atoms of amino acids that are predicted to influence remdesivir binding are shown as red spheres. The amino acid Cys812 located at the catalytic site is shown as green sphere. The location of Cα atoms of remdesivir resistance conferring amino acid Val556 is shown as blue sphere and labeled.

**Supplemental FIG 5** Distribution of subclades characterized by particular amino acid replacements in spike protein.

**Supplemental FIG 6** Biochemical characterization of single amino acid variants of spike protein RBD. (A, B) Size-exclusion chromatography (SEC) traces of the indicated spike-RBD variants. Dashed line indicates the elution peak of spike-6P. (C) Thermostability analysis of RBD variants. Each sample had three replicates and only mean values were plotted. Black vertical dashed line indicates the first melting temperature of 6P-D614G. (D) ELISA-based binding affinities for ACE2 and (E) neutralizing monoclonal antibody CR3022 to the indicated RBD variants.

## [Supplemental Table Legends]

**Supplemental Table 1** Patient demographics in wave 1 and wave 2.

**Supplemental Table 2** Classifier accuracy scores and performance of machine learning models.

**Supplemental Table 3** Pearson correlation coefficient data for correlation analysis.

**Supplemental Table 4** Primers and plasmids used for the *in vitro* characterization of recombinant proteins with single amino acid replacements in the receptor binding domain (RBD) region of spike protein, and their biophysical properties. To test the hypothesis that RBD amino acid changes enhance viral fitness, we expressed spike variants with the Asp614Gly replacement and 13 clinical RBD variants identified in our genome sequencing studies. Table S4A contains the primers used, Table S4B contains the plasmid construct information, and Table S4C contains the biophysical properties of the resultant spike protein variants.

## REFERENCES

1. 2020. World Health Organization Coronavirus Disease 2019 (COVID-19) Situation Report. https://www.who.int/docs/default-source/coronaviruse/situation-reports/20200420-sitrep-91-covid-19.pdf?sfvrsn=fcf0670b_4. Accessed April 21.

2. Gorbalenya AE, Baker SC, Baric RS, de Groot RJ, Drosten C, Gulyaeva AA, Haagmans BL, Lauber C, Leontovich AM, Neuman BW, Penzar D, Perlman S, Poon LLM, Samborskiy DV, Sidorov IA, Sola I, Ziebuhr J, Coronaviridae Study Group of the International Committee on Taxonomy of V. 2020. The species Severe acute respiratory syndrome-related coronavirus: classifying 2019-nCoV and naming it SARS-CoV-2. Nature Microbiology 5:536–544.

3. Wang C, Horby PW, Hayden FG, Gao GF. 2020. A novel coronavirus outbreak of global health concern. Lancet 395:470–473.

4. Perlman S. 2020. Another Decade, Another Coronavirus. New England Journal of Medicine 382:760–762.

5. Allel K, Tapia-Muñoz T, Morris W. 2020. Country-level factors associated with the early spread of COVID-19 cases at 5, 10 and 15 days since the onset. Glob Public Health doi:10.1080/17441692.2020.1814835:1-14.

6. Huang C, Wang Y, Li X, Ren L, Zhao J, Hu Y, Zhang L, Fan G, Xu J, Gu X, Cheng Z, Yu T, Xia J, Wei Y, Wu W, Xie X, Yin W, Li H, Liu M, Xiao Y, Gao H, Guo L, Xie J, Wang G, Jiang R, Gao Z, Jin Q, Wang J, Cao B. 2020. Clinical features of patients infected with 2019 novel coronavirus in Wuhan, China. Lancet 395:497–506.

7. Zhu N, Zhang D, Wang W, Li X, Yang B, Song J, Zhao X, Huang B, Shi W, Lu R, Niu P, Zhan F, Ma X, Wang D, Xu W, Wu G, Gao GF, Tan W. 2020. A Novel Coronavirus from Patients with Pneumonia in China, 2019. New England Journal of Medicine 382:727–733.

8. Chan JF, Yuan S, Kok KH, To KK, Chu H, Yang J, Xing F, Liu J, Yip CC, Poon RW, Tsoi HW, Lo SK, Chan KH, Poon VK, Chan WM, Ip JD, Cai JP, Cheng VC, Chen H, Hui CK, Yuen KY. 2020. A familial cluster of pneumonia associated with the 2019 novel coronavirus indicating person-to-person transmission: a study of a family cluster. Lancet 395:514–523.

9. Wu F, Zhao S, Yu B, Chen YM, Wang W, Song ZG, Hu Y, Tao ZW, Tian JH, Pei YY, Yuan ML, Zhang YL, Dai FH, Liu Y, Wang QM, Zheng JJ, Xu L, Holmes EC, Zhang YZ. 2020. A new coronavirus associated with human respiratory disease in China. Nature 579:265–269.

10. Bedford T, Greninger AL, Roychoudhury P, Starita LM, Famulare M, Huang M-L, Nalla A, Pepper G, Reinhardt A, Xie H, Shrestha L, Nguyen TN, Adler A, Brandstetter E, Cho S, Giroux D, Han PD, Fay K, Frazar CD, Ilcisin M, Lacombe K, Lee J, Kiavand A, Richardson M, Sibley TR, Truong M, Wolf CR, Nickerson DA, Rieder MJ, Englund JA, Hadfield J, Hodcroft EB, Huddleston J, Moncla LH, Müller NF, Neher RA, Deng X, Gu W, Federman S, Chiu C, Duchin J, Gautom R, Melly G, Hiatt B, Dykema P, Lindquist S, Queen K, Tao Y, Uehara A, Tong S, et al. 2020. Cryptic transmission of SARS-CoV-2 in Washington State. medRxiv doi:10.1101/2020.04.02.20051417:2020.04.02.20051417.

11. Gonzalez-Reiche AS, Hernandez MM, Sullivan MJ, Ciferri B, Alshammary H, Obla A, Fabre S, Kleiner G, Polanco J, Khan Z, Alburquerque B, van de Guchte A, Dutta J, Francoeur N, Melo BS, Oussenko I, Deikus G, Soto J, Sridhar SH, Wang Y-C, Twyman K, Kasarskis A, Altman DR, Smith M, Sebra R, Aberg J, Krammer F, García-Sastre A, Luksza M, Patel G, Paniz-Mondolfi A, Gitman M, Sordillo EM, Simon V, van Bakel H. 2020. Introductions and early spread of SARS-CoV-2 in the New York City area. Science 369:297–301.

12. Health N. 2020. COVID-19 Data. https://www1.nyc.gov/site/doh/covid/covid-19-data.page. Accessed August 19.

13. County K. 2020. Daily COVID-19 outbreak summary. https://www.kingcounty.gov/depts/health/covid-19/data/daily-summary.aspx. Accessed August 18.

14. Cline M, Emerson M, bratter j, howell j, Jeanty P. 2012. Houston Region Grows More Racially/Ethnically Diverse,With Small Declines in Segregation. A Joint Report Analyzing Census Data from 1990, 2000, and 2010.

15. Emerson M, Bratter J, Howell J, Jeanty P, Cline M. 2012. Houston Region Grows More Racially/Ethnically Diverse, With Small Declines in Segregation. A Joint Report Analyzing Census Data from 1990, 2000, and 2010. Kinder Institute for Urban Research & the Hobby Center for the Study of Texas,

16. Services THaH. 2020. Texas Health and Human Services. https://hhs.texas.gov/. Accessed August 18.

17. Vahidy FS, Drews AL, Masud FN, Schwartz RL, Askary BB, Boom ML, Phillips RA. 2020. Characteristics and Outcomes of COVID-19 Patients During Initial Peak and Resurgence in the Houston Metropolitan Area. Jama doi:10.1001/jama.2020.15301.

18. Diehl WE, Lin AE, Grubaugh ND, Carvalho LM, Kim K, Kyawe PP, McCauley SM, Donnard E, Kucukural A, McDonel P, Schaffner SF, Garber M, Rambaut A, Andersen KG, Sabeti PC, Luban J. 2016. Ebola Virus Glycoprotein with Increased Infectivity Dominated the 2013-2016 Epidemic. Cell 167:1088-1098.e6.

19. Urbanowicz RA, McClure CP, Sakuntabhai A, Sall AA, Kobinger G, Müller MA, Holmes EC, Rey FA, Simon-Loriere E, Ball JK. 2016. Human Adaptation of Ebola Virus during the West African Outbreak. Cell 167:1079-1087.e5.

20. Dietzel E, Schudt G, Krähling V, Matrosovich M, Becker S. 2017. Functional Characterization of Adaptive Mutations during the West African Ebola Virus Outbreak. J Virol 91.

21. Kachroo P, Eraso JM, Beres SB, Olsen RJ, Zhu L, Nasser W, Bernard PE, Cantu CC, Saavedra MO, Arredondo MJ, Strope B, Do H, Kumaraswami M, Vuopio J, Grondahl-Yli-Hannuksela K, Kristinsson KG, Gottfredsson M, Pesonen M, Pensar J, Davenport ER, Clark AG, Corander J, Caugant DA, Gaini S, Magnussen MD, Kubiak SL, Nguyen HAT, Long SW, Porter AR, DeLeo FR, Musser JM. 2019. Integrated analysis of population genomics, transcriptomics and virulence provides novel insights into Streptococcus pyogenes pathogenesis. Nat Genet 51:548–559.

22. Gao Y, Yan L, Huang Y, Liu F, Zhao Y, Cao L, Wang T, Sun Q, Ming Z, Zhang L, Ge J, Zheng L, Zhang Y, Wang H, Zhu Y, Zhu C, Hu T, Hua T, Zhang B, Yang X, Li J, Yang H, Liu Z, Xu W, Guddat LW, Wang Q, Lou Z, Rao Z. 2020. Structure of the RNA-dependent RNA polymerase from COVID-19 virus. Science doi:10.1126/science.abb7498:eabb7498.

23. Yin W, Mao C, Luan X, Shen D-D, Shen Q, Su H, Wang X, Zhou F, Zhao W, Gao M, Chang S, Xie Y-C, Tian G, Jiang H-W, Tao S-C, Shen J, Jiang Y, Jiang H, Xu Y, Zhang S, Zhang Y, Xu HE. 2020. Structural basis for inhibition of the RNA-dependent RNA polymerase from SARS-CoV-2 by remdesivir. Science 368:1499–1504.

24. Shannon A, Le NT, Selisko B, Eydoux C, Alvarez K, Guillemot JC, Decroly E, Peersen O, Ferron F, Canard B. 2020. Remdesivir and SARS-CoV-2: Structural requirements at both nsp12 RdRp and nsp14 Exonuclease active-sites. Antiviral Res 178:104793.

25. Gordon CJ, Tchesnokov EP, Woolner E, Perry JK, Feng JY, Porter DP, Gotte M. 2020. Remdesivir is a direct-acting antiviral that inhibits RNA-dependent RNA polymerase from severe acute respiratory syndrome coronavirus 2 with high potency. J Biol Chem doi:10.1074/jbc.RA120.013679.

26. Agostini ML, Andres EL, Sims AC, Graham RL, Sheahan TP, Lu X, Smith EC, Case JB, Feng JY, Jordan R, Ray AS, Cihlar T, Siegel D, Mackman RL, Clarke MO, Baric RS, Denison MR. 2018. Coronavirus Susceptibility to the Antiviral Remdesivir (GS-5734) Is Mediated by the Viral Polymerase and the Proofreading Exoribonuclease. mBio 9.

27. de Wit E, Feldmann F, Cronin J, Jordan R, Okumura A, Thomas T, Scott D, Cihlar T, Feldmann H. 2020. Prophylactic and therapeutic remdesivir (GS-5734) treatment in the rhesus macaque model of MERS-CoV infection. Proc Natl Acad Sci U S A 117:6771–6776.

28. Williamson BN, Feldmann F, Schwarz B, Meade-White K, Porter DP, Schulz J, van Doremalen N, Leighton I, Yinda CK, Pérez-Pérez L, Okumura A, Lovaglio J, Hanley PW, Saturday G, Bosio CM, Anzick S, Barbian K, Cihlar T, Martens C, Scott DP, Munster VJ, de Wit E. 2020. Clinical benefit of remdesivir in rhesus macaques infected with SARS-CoV-2. Nature 585:273–276.

29. Grein J, Ohmagari N, Shin D, Diaz G, Asperges E, Castagna A, Feldt T, Green G, Green ML, Lescure FX, Nicastri E, Oda R, Yo K, Quiros-Roldan E, Studemeister A, Redinski J, Ahmed S, Bernett J, Chelliah D, Chen D, Chihara S, Cohen SH, Cunningham J, D’Arminio Monforte A, Ismail S, Kato H, Lapadula G, L’Her E, Maeno T, Majumder S, Massari M, Mora-Rillo M, Mutoh Y, Nguyen D, Verweij E, Zoufaly A, Osinusi AO, DeZure A, Zhao Y, Zhong L, Chokkalingam A, Elboudwarej E, Telep L, Timbs L, Henne I, Sellers S, Cao H, Tan SK, Winterbourne L, Desai P, et al. 2020. Compassionate Use of Remdesivir for Patients with Severe Covid-19. N Engl J Med doi:10.1056/NEJMoa2007016.

30. Goldman JD, Lye DCB, Hui DS, Marks KM, Bruno R, Montejano R, Spinner CD, Galli M, Ahn MY, Nahass RG, Chen YS, SenGupta D, Hyland RH, Osinusi AO, Cao H, Blair C, Wei X, Gaggar A, Brainard DM, Towner WJ, Muñoz J, Mullane KM, Marty FM, Tashima KT, Diaz G, Subramanian A. 2020. Remdesivir for 5 or 10 Days in Patients with Severe Covid-19. N Engl J Med doi:10.1056/NEJMoa2015301.

31. Beigel JH, Tomashek KM, Dodd LE, Mehta AK, Zingman BS, Kalil AC, Hohmann E, Chu HY, Luetkemeyer A, Kline S, Lopez de Castilla D, Finberg RW, Dierberg K, Tapson V, Hsieh L, Patterson TF, Paredes R, Sweeney DA, Short WR, Touloumi G, Lye DC, Ohmagari N, Oh MD, Ruiz-Palacios GM, Benfield T, Fätkenheuer G, Kortepeter MG, Atmar RL, Creech CB, Lundgren J, Babiker AG, Pett S, Neaton JD, Burgess TH, Bonnett T, Green M, Makowski M, Osinusi A, Nayak S, Lane HC. 2020. Remdesivir for the Treatment of Covid-19 -Preliminary Report. N Engl J Med doi:10.1056/NEJMoa2007764.

32. Spinner CD, Gottlieb RL, Criner GJ, Arribas López JR, Cattelan AM, Soriano Viladomiu A, Ogbuagu O, Malhotra P, Mullane KM, Castagna A, Chai LYA, Roestenberg M, Tsang OTY, Bernasconi E, Le Turnier P, Chang SC, SenGupta D, Hyland RH, Osinusi AO, Cao H, Blair C, Wang H, Gaggar A, Brainard DM, McPhail MJ, Bhagani S, Ahn MY, Sanyal AJ, Huhn G, Marty FM. 2020. Effect of Remdesivir vs Standard Care on Clinical Status at 11 Days in Patients With Moderate COVID-19: A Randomized Clinical Trial. Jama doi:10.1001/jama.2020.16349.

33. Olender SA, Perez KK, Go AS, Balani B, Price-Haywood EG, Shah NS, Wang S, Walunas TL, Swaminathan S, Slim J, Chin B, De Wit S, Ali SM, Soriano Viladomiu A, Robinson P, Gottlieb RL, Tsang TYO, Lee IH, Haubrich RH, Chokkalingam AP, Lin L, Zhong L, Bekele BN, Mera-Giler R, Gallant J, Smith LE, Osinusi AO, Brainard DM, Hu H, Phulpin C, Edgar H, Diaz-Cuervo H, Bernardino JI. 2020. Remdesivir for Severe COVID-19 versus a Cohort Receiving Standard of Care. Clin Infect Dis doi:10.1093/cid/ciaa1041.

34. (CNCB) CNCfB. 2020. 2019 Novel Coronavirus Resource (2019nCoVR). https://bigd.big.ac.cn/ncov/about?lang=en. Accessed August 19.

35. Wrapp D, Wang N, Corbett KS, Goldsmith JA, Hsieh C-L, Abiona O, Graham BS, McLellan JS. 2020. Cryo-EM structure of the 2019-nCoV spike in the prefusion conformation. Science 367:1260–1263.

36. Walls AC, Park YJ, Tortorici MA, Wall A, McGuire AT, Veesler D. 2020. Structure, Function, and Antigenicity of the SARS-CoV-2 Spike Glycoprotein. Cell 181:281-292.e6.

37. Wang Q, Zhang Y, Wu L, Niu S, Song C, Zhang Z, Lu G, Qiao C, Hu Y, Yuen KY, Wang Q, Zhou H, Yan J, Qi J. 2020. Structural and Functional Basis of SARS-CoV-2 Entry by Using Human ACE2. Cell doi:10.1016/j.cell.2020.03.045.

38. Jackson LA, Anderson EJ, Rouphael NG, Roberts PC, Makhene M, Coler RN, McCullough MP, Chappell JD, Denison MR, Stevens LJ, Pruijssers AJ, McDermott A, Flach B, Doria-Rose NA, Corbett KS, Morabito KM, O’Dell S, Schmidt SD, Swanson PA, 2nd, Padilla M, Mascola JR, Neuzil KM, Bennett H, Sun W, Peters E, Makowski M, Albert J, Cross K, Buchanan W, Pikaart-Tautges R, Ledgerwood JE, Graham BS, Beigel JH. 2020. An mRNA Vaccine against SARS-CoV-2 -Preliminary Report. N Engl J Med doi:10.1056/NEJMoa2022483.

39. Folegatti PM, Ewer KJ, Aley PK, Angus B, Becker S, Belij-Rammerstorfer S, Bellamy D, Bibi S, Bittaye M, Clutterbuck EA, Dold C, Faust SN, Finn A, Flaxman AL, Hallis B, Heath P, Jenkin D, Lazarus R, Makinson R, Minassian AM, Pollock KM, Ramasamy M, Robinson H, Snape M, Tarrant R, Voysey M, Green C, Douglas AD, Hill AVS, Lambe T, Gilbert SC, Pollard AJ. 2020. Safety and immunogenicity of the ChAdOx1 nCoV-19 vaccine against SARS-CoV-2: a preliminary report of a phase 1/2, single-blind, randomised controlled trial. Lancet 396:467–478.

40. Zhu FC, Guan XH, Li YH, Huang JY, Jiang T, Hou LH, Li JX, Yang BF, Wang L, Wang WJ, Wu SP, Wang Z, Wu XH, Xu JJ, Zhang Z, Jia SY, Wang BS, Hu Y, Liu JJ, Zhang J, Qian XA, Li Q, Pan HX, Jiang HD, Deng P, Gou JB, Wang XW, Wang XH, Chen W. 2020. Immunogenicity and safety of a recombinant adenovirus type-5-vectored COVID-19 vaccine in healthy adults aged 18 years or older: a randomised, double-blind, placebo-controlled, phase 2 trial. Lancet 396:479–488.

41. Brouwer PJM, Caniels TG, van der Straten K, Snitselaar JL, Aldon Y, Bangaru S, Torres JL, Okba NMA, Claireaux M, Kerster G, Bentlage AEH, van Haaren MM, Guerra D, Burger JA, Schermer EE, Verheul KD, van der Velde N, van der Kooi A, van Schooten J, van Breemen MJ, Bijl TPL, Sliepen K, Aartse A, Derking R, Bontjer I, Kootstra NA, Wiersinga WJ, Vidarsson G, Haagmans BL, Ward AB, de Bree GJ, Sanders RW, van Gils MJ. 2020. Potent neutralizing antibodies from COVID-19 patients define multiple targets of vulnerability. Science 369:643–650.

42. Chi X, Yan R, Zhang J, Zhang G, Zhang Y, Hao M, Zhang Z, Fan P, Dong Y, Yang Y, Chen Z, Guo Y, Zhang J, Li Y, Song X, Chen Y, Xia L, Fu L, Hou L, Xu J, Yu C, Li J, Zhou Q, Chen W. 2020. A neutralizing human antibody binds to the N-terminal domain of the Spike protein of SARS-CoV-2. Science 369:650–655.

43. Wec AZ, Wrapp D, Herbert AS, Maurer DP, Haslwanter D, Sakharkar M, Jangra RK, Dieterle ME, Lilov A, Huang D, Tse LV, Johnson NV, Hsieh C-L, Wang N, Nett JH, Champney E, Burnina I, Brown M, Lin S, Sinclair M, Johnson C, Pudi S, Bortz R, Wirchnianski AS, Laudermilch E, Florez C, Fels JM, O’Brien CM, Graham BS, Nemazee D, Burton DR, Baric RS, Voss JE, Chandran K, Dye JM, McLellan JS, Walker LM. 2020. Broad neutralization of SARS-related viruses by human monoclonal antibodies. Science 369:731–736.

44. Zost SJ, Gilchuk P, Case JB, Binshtein E, Chen RE, Nkolola JP, Schäfer A, Reidy JX, Trivette A, Nargi RS, Sutton RE, Suryadevara N, Martinez DR, Williamson LE, Chen EC, Jones T, Day S, Myers L, Hassan AO, Kafai NM, Winkler ES, Fox JM, Shrihari S, Mueller BK, Meiler J, Chandrashekar A, Mercado NB, Steinhardt JJ, Ren K, Loo YM, Kallewaard NL, McCune BT, Keeler SP, Holtzman MJ, Barouch DH, Gralinski LE, Baric RS, Thackray LB, Diamond MS, Carnahan RH, Crowe JE, Jr. 2020. Potently neutralizing and protective human antibodies against SARS-CoV-2. Nature 584:443–449.

45. Greaney AJ, Starr TN, Gilchuk P, Zost SJ, Binshtein E, Loes AN, Hilton SK, Huddleston J, Eguia R, Crawford KH, Dingens AS, Nargi RS, Sutton RE, Suryadevara N, Rothlauf PW, Liu Z, Whelan SP, Carnahan RH, Crowe JE, Bloom JD. 2020. Complete mapping of mutations to the SARS-CoV-2 spike receptor-binding domain that escape antibody recognition. bioRxiv doi:10.1101/2020.09.10.292078:2020.09.10.292078.

46. Baum A, Copin R, Ajithdoss D, Zhou A, Lanza K, Negron N, Ni M, Wei Y, Atwal GS, Oyejide A, Goez-Gazi Y, Dutton J, Clemmons E, Staples HM, Bartley C, Klaffke B, Alfson K, Gazi M, Gonzales O, Dick E, Carrion R, Pessaint L, Porto M, Cook A, Brown R, Ali V, Greenhouse J, Taylor T, Andersen H, Lewis MG, Stahl N, Murphy AJ, Yancopoulos GD, Kyratsous CA. 2020. REGN-COV2 antibody cocktail prevents and treats SARS-CoV-2 infection in rhesus macaques and hamsters. bioRxiv doi:10.1101/2020.08.02.233320:2020.08.02.233320.

47. Baum A, Fulton BO, Wloga E, Copin R, Pascal KE, Russo V, Giordano S, Lanza K, Negron N, Ni M, Wei Y, Atwal GS, Murphy AJ, Stahl N, Yancopoulos GD, Kyratsous CA. 2020. Antibody cocktail to SARS-CoV-2 spike protein prevents rapid mutational escape seen with individual antibodies. Science 369:1014–1018.

48. Barnes CO, West AP, Jr., Huey-Tubman KE, Hoffmann MAG, Sharaf NG, Hoffman PR, Koranda N, Gristick HB, Gaebler C, Muecksch F, Lorenzi JCC, Finkin S, Hägglöf T, Hurley A, Millard KG, Weisblum Y, Schmidt F, Hatziioannou T, Bieniasz PD, Caskey M, Robbiani DF, Nussenzweig MC, Bjorkman PJ. 2020. Structures of Human Antibodies Bound to SARS-CoV-2 Spike Reveal Common Epitopes and Recurrent Features of Antibodies. Cell 182:828-842.e16.

49. Alsoussi WB, Turner JS, Case JB, Zhao H, Schmitz AJ, Zhou JQ, Chen RE, Lei T, Rizk AA, McIntire KM, Winkler ES, Fox JM, Kafai NM, Thackray LB, Hassan AO, Amanat F, Krammer F, Watson CT, Kleinstein SH, Fremont DH, Diamond MS, Ellebedy AH. 2020. A Potently Neutralizing Antibody Protects Mice against SARS-CoV-2 Infection. J Immunol 205:915–922.

50. Salazar E, Kuchipudi SV, Christensen PA, Eagar T, Yi X, Zhao P, Jin Z, Long SW, Olsen RJ, Chen J, Castillo B, Leveque C, Towers D, Lavinder JJ, Gollihar J, Cardona JA, Ippolito GC, Nissly RH, Bird I, Greenawalt D, Rossi RM, Gontu A, Srinivasan S, Poojary I, Cattadori IM, Hudson P, Josleyn NM, Prugar L, Huie KE, Herbert AS, Bernard DW, Dye JM, Kapur V, Musser JM. 2020. Convalescent plasma anti-SARS-CoV-2 spike protein ectodomain and receptor binding domain IgG correlate with virus neutralization. The Journal of Clinical Investigation doi:10.1172/JCI141206.

51. Salazar E, Christensen PA, Graviss EA, Nguyen DT, Castillo B, Chen J, Lopez BV, Eagar TN, Yi X, Zhao P, Rogers J, Shehabeldin A, Joseph D, Leveque C, Olsen RJ, Bernard DW, Gollihar J, Musser JM. 2020. Treatment of COVID-19 Patients with Convalescent Plasma Reveals a Signal of Significantly Decreased Mortality. Am J Pathol doi:10.1016/j.ajpath.2020.08.001.

52. Starr TN, Greaney AJ, Hilton SK, Ellis D, Crawford KHD, Dingens AS, Navarro MJ, Bowen JE, Tortorici MA, Walls AC, King NP, Veesler D, Bloom JD. 2020. Deep Mutational Scanning of SARS-CoV-2 Receptor Binding Domain Reveals Constraints on Folding and ACE2 Binding. Cell doi:10.1016/j.cell.2020.08.012.

53. Steffen TL, Stone ET, Hassert M, Geerling E, Grimberg BT, Espino AM, Pantoja P, Climent C, Hoft DF, George SL, Sariol CA, Pinto AK, Brien JD. 2020. The receptor binding domain of SARS-CoV-2 spike is the key target of neutralizing antibody in human polyclonal sera. bioRxiv doi:10.1101/2020.08.21.261727:2020.08.21.261727.

54. Corbett KS, Flynn B, Foulds KE, Francica JR, Boyoglu-Barnum S, Werner AP, Flach B, O’Connell S, Bock KW, Minai M, Nagata BM, Andersen H, Martinez DR, Noe AT, Douek N, Donaldson MM, Nji NN, Alvarado GS, Edwards DK, Flebbe DR, Lamb E, Doria-Rose NA, Lin BC, Louder MK, O’Dell S, Schmidt SD, Phung E, Chang LA, Yap C, Todd J-PM, Pessaint L, Van Ry A, Browne S, Greenhouse J, Putman-Taylor T, Strasbaugh A, Campbell T-A, Cook A, Dodson A, Steingrebe K, Shi W, Zhang Y, Abiona OM, Wang L, Pegu A, Yang ES, Leung K, Zhou T, Teng I-T, Widge A, et al. 2020. Evaluation of the mRNA-1273 Vaccine against SARS-CoV-2 in Nonhuman Primates. New England Journal of Medicine doi:10.1056/NEJMoa2024671.

55. van Doremalen N, Lambe T, Spencer A, Belij-Rammerstorfer S, Purushotham JN, Port JR, Avanzato VA, Bushmaker T, Flaxman A, Ulaszewska M, Feldmann F, Allen ER, Sharpe H, Schulz J, Holbrook M, Okumura A, Meade-White K, Pérez-Pérez L, Edwards NJ, Wright D, Bissett C, Gilbride C, Williamson BN, Rosenke R, Long D, Ishwarbhai A, Kailath R, Rose L, Morris S, Powers C, Lovaglio J, Hanley PW, Scott D, Saturday G, de Wit E, Gilbert SC, Munster VJ. 2020. ChAdOx1 nCoV-19 vaccine prevents SARS-CoV-2 pneumonia in rhesus macaques. Nature doi:10.1038/s41586-020-2608-y.

56. Wang C, Li W, Drabek D, Okba NMA, van Haperen R, Osterhaus A, van Kuppeveld FJM, Haagmans BL, Grosveld F, Bosch BJ. 2020. A human monoclonal antibody blocking SARS-CoV-2 infection. Nat Commun 11:2251.

57. Ju B, Zhang Q, Ge J, Wang R, Sun J, Ge X, Yu J, Shan S, Zhou B, Song S, Tang X, Yu J, Lan J, Yuan J, Wang H, Zhao J, Zhang S, Wang Y, Shi X, Liu L, Zhao J, Wang X, Zhang Z, Zhang L. 2020. Human neutralizing antibodies elicited by SARS-CoV-2 infection. Nature 584:115–119.

58. Liu L, Wang P, Nair MS, Yu J, Rapp M, Wang Q, Luo Y, Chan JF, Sahi V, Figueroa A, Guo XV, Cerutti G, Bimela J, Gorman J, Zhou T, Chen Z, Yuen KY, Kwong PD, Sodroski JG, Yin MT, Sheng Z, Huang Y, Shapiro L, Ho DD. 2020. Potent neutralizing antibodies against multiple epitopes on SARS-CoV-2 spike. Nature 584:450–456.

59. Rogers TF, Zhao F, Huang D, Beutler N, Burns A, He W-t, Limbo O, Smith C, Song G, Woehl J, Yang L, Abbott RK, Callaghan S, Garcia E, Hurtado J, Parren M, Peng L, Ramirez S, Ricketts J, Ricciardi MJ, Rawlings SA, Wu NC, Yuan M, Smith DM, Nemazee D, Teijaro JR, Voss JE, Wilson IA, Andrabi R, Briney B, Landais E, Sok D, Jardine JG, Burton DR. 2020. Isolation of potent SARS-CoV-2 neutralizing antibodies and protection from disease in a small animal model. Science 369:956–963.

60. Hassan AO, Case JB, Winkler ES, Thackray LB, Kafai NM, Bailey AL, McCune BT, Fox JM, Chen RE, Alsoussi WB, Turner JS, Schmitz AJ, Lei T, Shrihari S, Keeler SP, Fremont DH, Greco S, McCray PB, Jr., Perlman S, Holtzman MJ, Ellebedy AH, Diamond MS. 2020. A SARS-CoV-2 Infection Model in Mice Demonstrates Protection by Neutralizing Antibodies. Cell 182:744-753.e4.

61. Chandrashekar A, Liu J, Martinot AJ, McMahan K, Mercado NB, Peter L, Tostanoski LH, Yu J, Maliga Z, Nekorchuk M, Busman-Sahay K, Terry M, Wrijil LM, Ducat S, Martinez DR, Atyeo C, Fischinger S, Burke JS, Slein MD, Pessaint L, Van Ry A, Greenhouse J, Taylor T, Blade K, Cook A, Finneyfrock B, Brown R, Teow E, Velasco J, Zahn R, Wegmann F, Abbink P, Bondzie EA, Dagotto G, Gebre MS, He X, Jacob-Dolan C, Kordana N, Li Z, Lifton MA, Mahrokhian SH, Maxfield LF, Nityanandam R, Nkolola JP, Schmidt AG, Miller AD, Baric RS, Alter G, Sorger PK, Estes JD, et al. 2020. SARS-CoV-2 infection protects against rechallenge in rhesus macaques. Science 369:812–817.

62. Mercado NB, Zahn R, Wegmann F, Loos C, Chandrashekar A, Yu J, Liu J, Peter L, McMahan K, Tostanoski LH, He X, Martinez DR, Rutten L, Bos R, van Manen D, Vellinga J, Custers J, Langedijk JP, Kwaks T, Bakkers MJG, Zuijdgeest D, Huber SKR, Atyeo C, Fischinger S, Burke JS, Feldman J, Hauser BM, Caradonna TM, Bondzie EA, Dagotto G, Gebre MS, Hoffman E, Jacob-Dolan C, Kirilova M, Li Z, Lin Z, Mahrokhian SH, Maxfield LF, Nampanya F, Nityanandam R, Nkolola JP, Patel S, Ventura JD, Verrington K, Wan H, Pessaint L, Ry AV, Blade K, Strasbaugh A, Cabus M, et al. 2020. Single-shot Ad26 vaccine protects against SARS-CoV-2 in rhesus macaques. Nature doi:10.1038/s41586-020-2607-z.

63. Yuan M, Wu NC, Zhu X, Lee CD, So RTY, Lv H, Mok CKP, Wilson IA. 2020. A highly conserved cryptic epitope in the receptor binding domains of SARS-CoV-2 and SARS-CoV. Science 368:630–633.

64. Hsieh C-L, Goldsmith JA, Schaub JM, DiVenere AM, Kuo H-C, Javanmardi K, Le KC, Wrapp D, Lee AG, Liu Y, Chou C-W, Byrne PO, Hjorth CK, Johnson NV, Ludes-Meyers J, Nguyen AW, Park J, Wang N, Amengor D, Lavinder JJ, Ippolito GC, Maynard JA, Finkelstein IJ, McLellan JS. 2020. Structure-based design of prefusion-stabilized SARS-CoV-2 spikes. Science 369:1501–1505.

65. Woo H, Park SJ, Choi YK, Park T, Tanveer M, Cao Y, Kern NR, Lee J, Yeom MS, Croll TI, Seok C, Im W. 2020. Developing a Fully Glycosylated Full-Length SARS-CoV-2 Spike Protein Model in a Viral Membrane. J Phys Chem B 124:7128–7137.

66. Korber B, Fischer WM, Gnanakaran S, Yoon H, Theiler J, Abfalterer W, Hengartner N, Giorgi EE, Bhattacharya T, Foley B, Hastie KM, Parker MD, Partridge DG, Evans CM, Freeman TM, de Silva TI, McDanal C, Perez LG, Tang H, Moon-Walker A, Whelan SP, LaBranche CC, Saphire EO, Montefiori DC. 2020. Tracking Changes in SARS-CoV-2 Spike: Evidence that D614G Increases Infectivity of the COVID-19 Virus. Cell 182:812-827.e19.

67. Hu J, He C-L, Gao Q-Z, Zhang G-J, Cao X-X, Long Q-X, Deng H-J, Huang L-Y, Chen J, Wang K, Tang N, Huang A-L. 2020. D614G mutation of SARS-CoV-2 spike protein enhances viral infectivity. bioRxiv doi:10.1101/2020.06.20.161323:2020.06.20.161323.

68. Lorenzo-Redondo R, Nam HH, Roberts SC, Simons LM, Jennings LJ, Qi C, Achenbach CJ, Hauser AR, Ison MG, Hultquist JF, Ozer EA. 2020. A Unique Clade of SARS-CoV-2 Viruses is Associated with Lower Viral Loads in Patient Upper Airways. medRxiv doi:10.1101/2020.05.19.20107144:2020.05.19.20107144.

69. Cassia Wagner PR, Chris D. Frazar, Jover Lee, Nicola F. Müller, Louise H. Moncla, James Hadfield, Emma B. Hodcroft, Benjamin Pelle, Matthew Richardson, Caitlin Behrens, Meei-Li Huang, Patrick Mathias, Gregory Pepper, Lasata Shrestha, Hong Xie, Amin Addetia, Truong Nguyen, Victoria M Rachleff, Romesh Gautom, Geoff Melly, Brian Hiatt, Philip Dykema, Amanda Adler, Elisabeth Brandstetter, Peter D. Han, Kairsten Fay, Misja Ilcisin, Kirsten Lacombe, Thomas R. Sibley, Melissa Truong, Caitlin R. Wolf, Karen Cowgill, Stephanie Schrag, Jeff Duchin, Michael Boeckh, Janet A. Englund, Michael Famulare, Barry R. Lutz, Mark J. Rieder, Matthew Thompson, Richard A. Neher, Geoffrey S. Baird, Lea M. Starita, Helen Y. Chu, Jay Shendure, Scott Lindquist, Deborah A. Nickerson, Alexander L. Greninger, Keith R. Jerome, Trevor Bedford. 2020. Comparing viral load and clinical outcomes in Washington State across D614G substitution in spike protein of SARS-CoV-2. https://github.com/blab/ncov-wa-d614g. Accessed September 8.

70. Volz EM, Hill V, McCrone JT, Price A, Jorgensen D, O’Toole A, Southgate JA, Johnson R, Jackson B, Nascimento FF, Rey SM, Nicholls SM, Colquhoun RM, da Silva Filipe A, Shepherd JG, Pascall DJ, Shah R, Jesudason N, Li K, Jarrett R, Pacchiarini N, Bull M, Geidelberg L, Siveroni I, Goodfellow IG, Loman NJ, Pybus O, Robertson DL, Thomson EC, Rambaut A, Connor TR. 2020. Evaluating the effects of SARS-CoV-2 Spike mutation D614G on transmissibility and pathogenicity. medRxiv doi:10.1101/2020.07.31.20166082:2020.07.31.20166082.

71. Lv Z, Deng Y-Q, Ye Q, Cao L, Sun C-Y, Fan C, Huang W, Sun S, Sun Y, Zhu L, Chen Q, Wang N, Nie J, Cui Z, Zhu D, Shaw N, Li X-F, Li Q, Xie L, Wang Y, Rao Z, Qin C-F, Wang X. 2020. Structural basis for neutralization of SARS-CoV-2 and SARS-CoV by a potent therapeutic antibody. Science 369:1505–1509.

72. Yurkovetskiy L, Wang X, Pascal KE, Tomkins-Tinch C, Nyalile T, Wang Y, Baum A, Diehl WE, Dauphin A, Carbone C, Veinotte K, Egri SB, Schaffner SF, Lemieux JE, Munro J, Rafique A, Barve A, Sabeti PC, Kyratsous CA, Dudkina N, Shen K, Luban J. 2020. Structural and Functional Analysis of the D614G SARS-CoV-2 Spike Protein Variant. bioRxiv doi:10.1101/2020.07.04.187757:2020.07.04.187757.

73. Zhang L, Jackson CB, Mou H, Ojha A, Rangarajan ES, Izard T, Farzan M, Choe H. 2020. The D614G mutation in the SARS-CoV-2 spike protein reduces S1 shedding and increases infectivity. bioRxiv doi:10.1101/2020.06.12.148726:2020.06.12.148726.

74. Huo J, Zhao Y, Ren J, Zhou D, Duyvesteyn HME, Ginn HM, Carrique L, Malinauskas T, Ruza RR, Shah PNM, Tan TK, Rijal P, Coombes N, Bewley KR, Tree JA, Radecke J, Paterson NG, Supasa P, Mongkolsapaya J, Screaton GR, Carroll M, Townsend A, Fry EE, Owens RJ, Stuart DI. 2020. Neutralization of SARS-CoV-2 by Destruction of the Prefusion Spike. Cell Host Microbe doi:10.1016/j.chom.2020.06.010.

75. Long SW, Olsen RJ, Christensen PA, Bernard DW, Davis JR, Shukla M, Nguyen M, Ojeda Saavedra M, Cantu CC, Yerramilli P, Pruitt L, Subedi S, Hendrickson H, Eskandari G, Kumaraswami M, McLellan JS, Musser JM. 2020. Molecular Architecture of Early Dissemination and Evolution of the SARS-CoV-2 Virus in Metropolitan Houston, Texas. bioRxiv doi:10.1101/2020.05.01.072652:2020.05.01.072652.

76. Plante JA, Liu Y, Liu J, Xia H, Johnson BA, Lokugamage KG, Zhang X, Muruato AE, Zou J, Fontes-Garfias CR, Mirchandani D, Scharton D, Bilello JP, Ku Z, An Z, Kalveram B, Freiberg AN, Menachery VD, Xie X, Plante KS, Weaver SC, Shi P-Y. 2020. Spike mutation D614G alters SARS-CoV-2 fitness and neutralization susceptibility. bioRxiv doi:10.1101/2020.09.01.278689:2020.09.01.278689.

77. Latz CA, DeCarlo C, Boitano L, Png CYM, Patell R, Conrad MF, Eagleton M, Dua A. 2020. Blood type and outcomes in patients with COVID-19. Ann Hematol 99:2113–2118.

78. Wu BB, Gu DZ, Yu JN, Yang J, Shen WQ. 2020. Association between ABO blood groups and COVID-19 infection, severity and demise: A systematic review and meta-analysis. Infect Genet Evol 84:104485.

79. Zhao J, Yang Y, Huang H, Li D, Gu D, Lu X, Zhang Z, Liu L, Liu T, Liu Y, He Y, Sun B, Wei M, Yang G, Wang X, Zhang L, Zhou X, Xing M, Wang PG. 2020. Relationship between the ABO Blood Group and the COVID-19 Susceptibility. Clin Infect Dis doi:10.1093/cid/ciaa1150.

80. Zietz M, Tatonetti NP. 2020. Testing the association between blood type and COVID-19 infection, intubation, and death. medRxiv doi:10.1101/2020.04.08.20058073.

81. Lemieux J, Siddle KJ, Shaw BM, Loreth C, Schaffner S, Gladden-Young A, Adams G, Fink T, Tomkins-Tinch CH, Krasilnikova LA, Deruff KC, Rudy M, Bauer MR, Lagerborg KA, Normandin E, Chapman SB, Reilly SK, Anahtar MN, Lin AE, Carter A, Myhrvold C, Kemball M, Chaluvadi S, Cusick C, Flowers K, Neumann A, Cerrato F, Farhat M, Slater D, Harris JB, Branda J, Hooper D, Gaeta JM, Baggett TP, O’Connell J, Gnirke A, Lieberman TD, Philippakis A, Burns M, Brown C, Luban J, Ryan ET, Turbett SE, LaRocque RC, Hanage WP, Gallagher G, Madoff LC, Smole S, Pierce VM, Rosenberg ES, et al. 2020. Phylogenetic analysis of SARS-CoV-2 in the Boston area highlights the role of recurrent importation and superspreading events. medRxiv doi:10.1101/2020.08.23.20178236:2020.08.23.20178236.

82. Gudbjartsson DF, Helgason A, Jonsson H, Magnusson OT, Melsted P, Norddahl GL, Saemundsdottir J, Sigurdsson A, Sulem P, Agustsdottir AB, Eiriksdottir B, Fridriksdottir R, Gardarsdottir EE, Georgsson G, Gretarsdottir OS, Gudmundsson KR, Gunnarsdottir TR, Gylfason A, Holm H, Jensson BO, Jonasdottir A, Jonsson F, Josefsdottir KS, Kristjansson T, Magnusdottir DN, le Roux L, Sigmundsdottir G, Sveinbjornsson G, Sveinsdottir KE, Sveinsdottir M, Thorarensen EA, Thorbjornsson B, Löve A, Masson G, Jonsdottir I, Möller AD, Gudnason T, Kristinsson KG, Thorsteinsdottir U, Stefansson K. 2020. Spread of SARS-CoV-2 in the Icelandic Population. N Engl J Med doi:10.1056/NEJMoa2006100.

83. Candido DS, Claro IM, de Jesus JG, Souza WM, Moreira FRR, Dellicour S, Mellan TA, du Plessis L, Pereira RHM, Sales FCS, Manuli ER, Thézé J, Almeida L, Menezes MT, Voloch CM, Fumagalli MJ, Coletti TM, da Silva CAM, Ramundo MS, Amorim MR, Hoeltgebaum HH, Mishra S, Gill MS, Carvalho LM, Buss LF, Prete CA, Ashworth J, Nakaya HI, Peixoto PS, Brady OJ, Nicholls SM, Tanuri A, Rossi ÁD, Braga CKV, Gerber AL, de C. Guimarães AP, Gaburo N, Alencar CS, Ferreira ACS, Lima CX, Levi JE, Granato C, Ferreira GM, Francisco RS, Granja F, Garcia MT, Moretti ML, Perroud MW, Castiñeiras TMPP, Lazari CS, et al. 2020. Evolution and epidemic spread of SARS-CoV-2 in Brazil. Science 369:1255–1260.

84. Ellinghaus D, Degenhardt F, Bujanda L, Buti M, Albillos A, Invernizzi P, Fernández J, Prati D, Baselli G, Asselta R, Grimsrud MM, Milani C, Aziz F, Kässens J, May S, Wendorff M, Wienbrandt L, Uellendahl-Werth F, Zheng T, Yi X, de Pablo R, Chercoles AG, Palom A, Garcia-Fernandez AE, Rodriguez-Frias F, Zanella A, Bandera A, Protti A, Aghemo A, Lleo A, Biondi A, Caballero-Garralda A, Gori A, Tanck A, Carreras Nolla A, Latiano A, Fracanzani AL, Peschuck A, Julià A, Pesenti A, Voza A, Jiménez D, Mateos B, Nafria Jimenez B, Quereda C, Paccapelo C, Gassner C, Angelini C, Cea C, Solier A, et al. 2020. Genomewide Association Study of Severe Covid-19 with Respiratory Failure. N Engl J Med doi:10.1056/NEJMoa2020283.

85. 2020. The COVID-19 Host Genetics Initiative, a global initiative to elucidate the role of host genetic factors in susceptibility and severity of the SARS-CoV-2 virus pandemic. Eur J Hum Genet 28:715–718.

86. Weisblum Y, Schmidt F, Zhang F, DaSilva J, Poston D, Lorenzi JCC, Muecksch F, Rutkowska M, Hoffmann H-H, Michailidis E, Gaebler C, Agudelo M, Cho A, Wang Z, Gazumyan A, Cipolla M, Luchsinger L, Hillyer CD, Caskey M, Robbiani DF, Rice CM, Nussenzweig MC, Hatziioannou T, Bieniasz PD. 2020. Escape from neutralizing antibodies by SARS-CoV-2 spike protein variants. bioRxiv doi:10.1101/2020.07.21.214759:2020.07.21.214759.

87. Li T, Han X, Wang Y, Gu C, Wang J, Hu C, Li S, Wang K, Luo F, Huang J, Long Y, Song S, Wang W, Hu J, Wu R, Mu S, Hao Y, Chen Q, Gao F, Shen M, Long S, Gong F, Li L, Wu Y, Xu W, Cai X, Qu D, Yuan Z, Gao Q, Zhang G, He C, Nai Y, Deng K, Du L, Tang N, Xie Y, Huang A, Jin A. 2020. A key linear epitope for a potent neutralizing antibody to SARS-CoV-2 S-RBD. bioRxiv doi:10.1101/2020.09.11.292631:2020.09.11.292631.

88. Hansen J, Baum A, Pascal KE, Russo V, Giordano S, Wloga E, Fulton BO, Yan Y, Koon K, Patel K, Chung KM, Hermann A, Ullman E, Cruz J, Rafique A, Huang T, Fairhurst J, Libertiny C, Malbec M, Lee W-y, Welsh R, Farr G, Pennington S, Deshpande D, Cheng J, Watty A, Bouffard P, Babb R, Levenkova N, Chen C, Zhang B, Romero Hernandez A, Saotome K, Zhou Y, Franklin M, Sivapalasingam S, Lye DC, Weston S, Logue J, Haupt R, Frieman M, Chen G, Olson W, Murphy AJ, Stahl N, Yancopoulos GD, Kyratsous CA. 2020. Studies in humanized mice and convalescent humans yield a SARS-CoV-2 antibody cocktail. Science 369:1010–1014.

89. Long SW, Olsen RJ, Eagar TN, Beres SB, Zhao P, Davis JJ, Brettin T, Xia F, Musser JM. 2017. Population Genomic Analysis of 1,777 Extended-Spectrum Beta-Lactamase-Producing <em>Klebsiella pneumoniae</em> Isolates, Houston, Texas: Unexpected Abundance of Clonal Group 307. mBio 8:e00489–17.

90. Stucker KM, Schobel SA, Olsen RJ, Hodges HL, Lin X, Halpin RA, Fedorova N, Stockwell TB, Tovchigrechko A, Das SR, Wentworth DE, Musser JM. 2015. Haemagglutinin mutations and glycosylation changes shaped the 2012/13 influenza A(H3N2) epidemic, Houston, Texas. Euro Surveill 20.

91. Katoh K, Standley DM. 2013. MAFFT multiple sequence alignment software version 7: improvements in performance and usability. Mol Biol Evol 30:772–80.

92. Waterhouse AM, Procter JB, Martin DM, Clamp M, Barton GJ. 2009. Jalview Version 2--a multiple sequence alignment editor and analysis workbench. Bioinformatics 25:1189–91.

93. Price MN, Dehal PS, Arkin AP. 2010. FastTree 2--approximately maximum-likelihood trees for large alignments. PLoS One 5:e9490.

94. Chen T, Guestrin C. 2016. XGBoost: A Scalable Tree Boosting System, abstr Proceedings of the 22nd ACM SIGKDD International Conference on Knowledge Discovery and Data Mining, San Francisco, California, USA, Association for Computing Machinery,

95. Nguyen M, Brettin T, Long SW, Musser JM, Olsen RJ, Olson R, Shukla M, Stevens RL, Xia F, Yoo H. 2018. Developing an in silico minimum inhibitory concentration panel test for Klebsiella pneumoniae. Scientific reports 8:421.

96. Nguyen M, Long SW, McDermott PF, Olsen RJ, Olson R, Stevens RL, Tyson GH, Zhao S, Davis JJ. 2019. Using machine learning to predict antimicrobial MICs and associated genomic features for nontyphoidal Salmonella. Journal of Clinical Microbiology 57:e01260–18.

97. Pedregosa F, Varoquaux, G., Gramfort, A., Michel, V., Thirion, B., Grisel, O., Blondel, M., Prettenhofer, P., Weiss, R., Dubourg, V., Vanderplas, J., Passos, A., Cournapeau, D., Brucher, M., Perrot, M., and Duchesnay, E. 2011. Scikit-learn: Machine Learning in Python. Journal of Machine Learning Research 12 (2011) 2825-2830.

98. Cai Y, Zhang J, Xiao T, Peng H, Sterling SM, Walsh RM, Jr., Rawson S, Rits-Volloch S, Chen B. 2020. Distinct conformational states of SARS-CoV-2 spike protein. Science doi:10.1126/science.abd4251.

